# Stress Biomarkers and Child Development in Young Children in Bangladesh

**DOI:** 10.1101/2023.09.12.23295429

**Authors:** Zachary Butzin-Dozier, Andrew N. Mertens, Sophia T. Tan, Douglas A. Granger, Helen O. Pitchik, Dora Il’yasova, Fahmida Tofail, Md. Ziaur Rahman, Ivan Spasojevic, Idan Shalev, Shahjahan Ali, Mohammed Rabiul Karim, Sunny Shahriar, Syeda Luthfa Famida, Gabrielle Shuman, Abul K. Shoab, Salma Akther, Md. Saheen Hossen, Palash Mutsuddi, Mahbubur Rahman, Leanne Unicomb, Kishor K. Das, Liying Yan, Ann Meyer, Christine P. Stewart, Alan Hubbard, Ruchira Tabassum Naved, Kausar Parvin, Md. Mahfuz Al Mamun, Stephen P. Luby, John M. Colford, Lia C. H. Fernald, Audrie Lin

**Author notes:** Contributed equally to this work (Lia C. H. Fernald, Audrie Lin). Corresponding author: Audrie Lin, PhD, MPH Assistant Professor Department of Microbiology and Environmental Toxicology University of California, Santa Cruz, 1156 High St, Santa Cruz, CA 95064.

## Abstract

**Background:** Hundreds of millions of children in low-and middle-income countries are exposed to chronic stressors, such as poverty, poor sanitation and hygiene, and sub-optimal nutrition. These stressors can have physiological consequences for children and may ultimately have detrimental effects on child development. This study explores associations between biological measures of chronic stress in early life and developmental outcomes in a large cohort of young children living in rural Bangladesh.

**Methods:** We assessed physiologic measures of stress in the first two years of life using measures of the hypothalamic-pituitary-adrenal (HPA) axis (salivary cortisol and glucocorticoid receptor gene methylation), the sympathetic-adrenal-medullary (SAM) system (salivary alpha-amylase, heart rate, and blood pressure), and oxidative status (F2-isoprostanes). We assessed child development in the first two years of life with the MacArthur-Bates Communicative Development Inventories (CDI), the WHO gross motor milestones, and the Extended Ages and Stages Questionnaire (EASQ). We compared development outcomes of children at the 75th and 25th percentiles of stress biomarker distributions while adjusting for potential confounders (hereafter referred to as contrasts) using generalized additive models, which are statistical models where the outcome is predicted by a potentially non-linear function of predictor variables.

**Results:** We analyzed data from 684 children (49% female) at both 14 and 28 months of age; we included an additional 765 children at 28 months of age. We observed 135 primary contrasts of the differences in child development outcomes at the 75^th^ and 25^th^ percentiles of stress biomarkers, where we detected significant relationships in 5 out of 30 contrasts (17%) of HPA axis activity, 1 out of 30 contrasts (3%) of SAM activity, and 3 out of 75 contrasts (4%) of oxidative status. These findings revealed that measures of HPA axis activity were associated with poor development outcomes. We did not find consistent evidence that markers of SAM system activity or oxidative status were associated with developmental status.

**Conclusions:** Our observations reveal associations between the physiological evidence of stress in the HPA axis with developmental status in early childhood. These findings add to the existing evidence exploring the developmental consequences of early life stress.

## INTRODUCTION

There are more than 250 million children in low and middle-income countries who are at risk of failing to reach their developmental potential (Black et al., 2017). Chronic stress experienced in early childhood can increase risk for poor developmental outcomes later in life (Franke, 2014; Grantham-McGregor et al., 2007; McEwen, 2011). Chronic early-life stress has the potential to negatively affect multiple biological systems and interfere with learning and memory, dysregulate metabolism and sleep, and negatively affect mental health (Lu et al., 2016; National Scientific Council on the Developing Child, 2014). Many children are exposed to stressors, such as iodine deficiency, inadequate cognitive stimulation, and infectious disease (e.g., malaria, diarrhea), that are associated with developmental delay (Rana et al., 2022; Walker et al., 2011, 2007). Stress biomarkers may reflect a mechanism by which stressors contribute to subsequent developmental status. An improved understanding of how specific biomarkers are related to development may have implications on pediatrics as well as policy (Shonkoff et al., 2022). Evaluating these associations in early childhood is particularly important, as this is an effective time to intervene on developmental outcomes (Engle et al., 2011).

Children in low-income, rural communities face many biological and psychosocial stressors (Walker et al., 2007). Social and economic factors can lead to developmental differences in children, and developmental neuroscience demonstrates how early experiences can influence development (Engle et al., 2011). Poverty and other sociocultural factors can alter neurological functioning, brain structure, and child behavior, which can in turn affect developmental status (Walker et al., 2007). Inadequate cognitive stimulation is a major risk factor for poor developmental outcomes, and children from low-income homes are less likely to have high quality stimulation at home (Engle et al., 2011; Walker et al., 2007). Although cumulative exposure to stressors leads to an increased risk of poor developmental outcomes, the impact of these exposures depends on their timing, co-occurrence, and an individual’s reactivity (physiologic response) to these stressors (Walker et al., 2011). These cumulative stressors are often measured through stress hormones (Engle et al., 2007).

Stress can be an adaptive or maladaptive response to challenging stimuli. Primary stress biochemical pathways include the hypothalamic-pituitary-adrenal (HPA) and sympathetic adreno-medullary (SAM) axes (Condon, 2018; Johnson et al., 2013). The first major component of the stress responses is the HPA axis which is controlled by a negative feedback loop in which pro-inflammatory cytokines stimulate HPA activation, triggering the release of anti-inflammatory cortisol, a glucocorticoid, which in turn dampens HPA axis activity (Johnson et al., 2013). The cortisol response enables individuals to respond to challenging circumstances, and cortisol is involved in mobilizing biological resources for metabolic, sensory, and learning processes (Booth et al., 2000). The *NR3C1* gene encodes glucocorticoid receptors, and early life stress is associated with *NR3C1* methylation (van der Knaap et al., 2014). Prolonged activation of the HPA axis and excess cortisol can lead to oxidative stress, which is an excess of reactive oxygen species relative to antioxidants (Aschbacher et al., 2013). Urinary F2-isoprostanes are the biomarkers most frequently used to measure oxidative status (Il’yasova et al., 2010; Montuschi et al., 2004; Pizzino et al., 2017). While oxidative stress can be harmful, reactive oxygen species play a critical role in the human body and immune function (Pizzino et al., 2017).

The second major component of the psychobiology of the stress response is activation of the SAM axis, which increases blood pressure and heart rate through the release of epinephrine and norepinephrine (Brietzke et al., 2012; McEwen, 2000). Whereas the HPA response to stress is linked with negative affect, distress, withdrawal, and being overwhelmed, the SAM response to stress is associated with increased engagement, cognitive effort, attentional focus, work, and arousal (Brietzke et al., 2012; McEwen, 2000). This axis also triggers the secretion of salivary alpha-amylase, a carbohydrate digestion enzyme that has been recently used as a salivary stress biomarker (Condon, 2018).

Researchers have evaluated individual differences in the psychobiology of the stress response and the consequences of these differences on early child development (Coe et al., 2003; Gunnar and Fisher, 2006; Kagan et al., 1987; Loman et al., 2010). Investigators have extended these research questions to assess the impact of various potential sources of stress including rural poverty (Fernald and Gunnar, 2009), maternal experience of intimate partner violence (Huth-Bocks et al., 2002; Levendosky et al., 2013), extreme neglect (Gunnar and Fisher, 2006; Reilly and Gunnar, 2019), parental divorce (Afifi et al., 2009), poor nutrition (Fernald et al., 2003; Fernald and Grantham-McGregor, 1998), and maternal substance use (Labella et al., 2021) on developmental outcomes in early childhood, although further research is needed to understand the mechanistic pathways through which these stressors can lead to developmental consequences. Correlational studies have indicated that poverty is associated with increased child cortisol, and a 2009 quasi-experimental study found that children from families who participated in a cash transfer program had lower cortisol compared to children from families who did not participate (Fernald and Gunnar, 2009; Schmidt et al., 2021). A 2003 study in Nepal as well as a 1998 study in Jamaica found that stress reactivity (salivary cortisol and heart rate) was associated with growth impairment (Fernald et al., 2003; Fernald and Grantham-McGregor, 1998).

Throughout decades of research on stress and development, several themes have emerged. First, the biobehavioral manifestations of chronic stress are heterogenous based on individual, familial, and community-level factors (Godoy et al., 2018). Second, differences in biological responses to stress (i.e. reactivity) are largely responsible for translating experiences into differential outcomes (Amstadter et al., 2016; Jiang et al., 2019; Schmidt et al., 2021; Wolf et al., 2018). Third, the social context of the family and quality of family care, including stimulating and nurturing behaviors, moderate the effects of exposures on development (Asok et al., 2013; Bernard et al., 2015; Booth et al., 2000; Fisher et al., 2007). Further evaluation of these associations in the context of low-and middle-income countries, where there is a large burden of both early-life chronic stress and poor developmental outcomes, as well as frequent exposure to inflammation and infection, may provide important insights in a high-risk population (Black et al., 2017).

This study aims to evaluate the associations between markers of HPA axis activity, SAM axis activity, oxidative stress, and child development outcomes in a cohort of young children (assessed at median ages 14 and 28 months) in rural Bangladesh. We hypothesized that decreased oxidative status and decreased salivary alpha-amylase would be associated with higher child development scores, while higher salivary cortisol, higher glucocorticoid receptor methylation, and higher heart rate and blood pressure would be associated with higher child development scores.

## METHODS AND MATERIALS

These analyses utilize data from the WASH Benefits study, described in detail previously (Luby et al., 2018; Tofail et al., 2018). The trial enrolled pregnant mothers in Bangladesh in their first or second trimester of pregnancy in rural subdistricts in Gazipur, Mymensingh, Tangail, and Kishoreganj and followed the cohort of children from birth until 2.5 years of age. Here, we describe observational analyses of the associations between child stress biomarkers and concurrent and subsequent child development in a subsample of children from the trial. Sample household enrollment characteristics remained similar to the main trial (Lin et al., 2020). This sample included 684 children aged 14 months (median age, Year 1) and 1,449 children (49% female) aged 28 months (median age, Year 2).

### Correlates of Stress Biomarkers

Child stress biomarkers included markers of the HPA axis, the SAM axis, and oxidative status. HPA axis biomarkers were salivary assessments of cortisol and glucocorticoid receptor (*NR3C1*) methylation. SAM axis measures were salivary alpha-amylase, resting heart rate, and mean arterial pressure. We measured oxidative status using four urinary F2-isoprostanes. We measured oxidative status markers at Year 1 (median age 14 months), while we assessed HPA and SAM axis biomarkers at Year 2 (median age 28 months; Table 1). We assessed resting heart rate and mean arterial pressure in four study arms, but we only measured other stress biomarker exposures in two arms, which led to a greater sample size at Year 2 compared to Year 1.

**Table 1.**
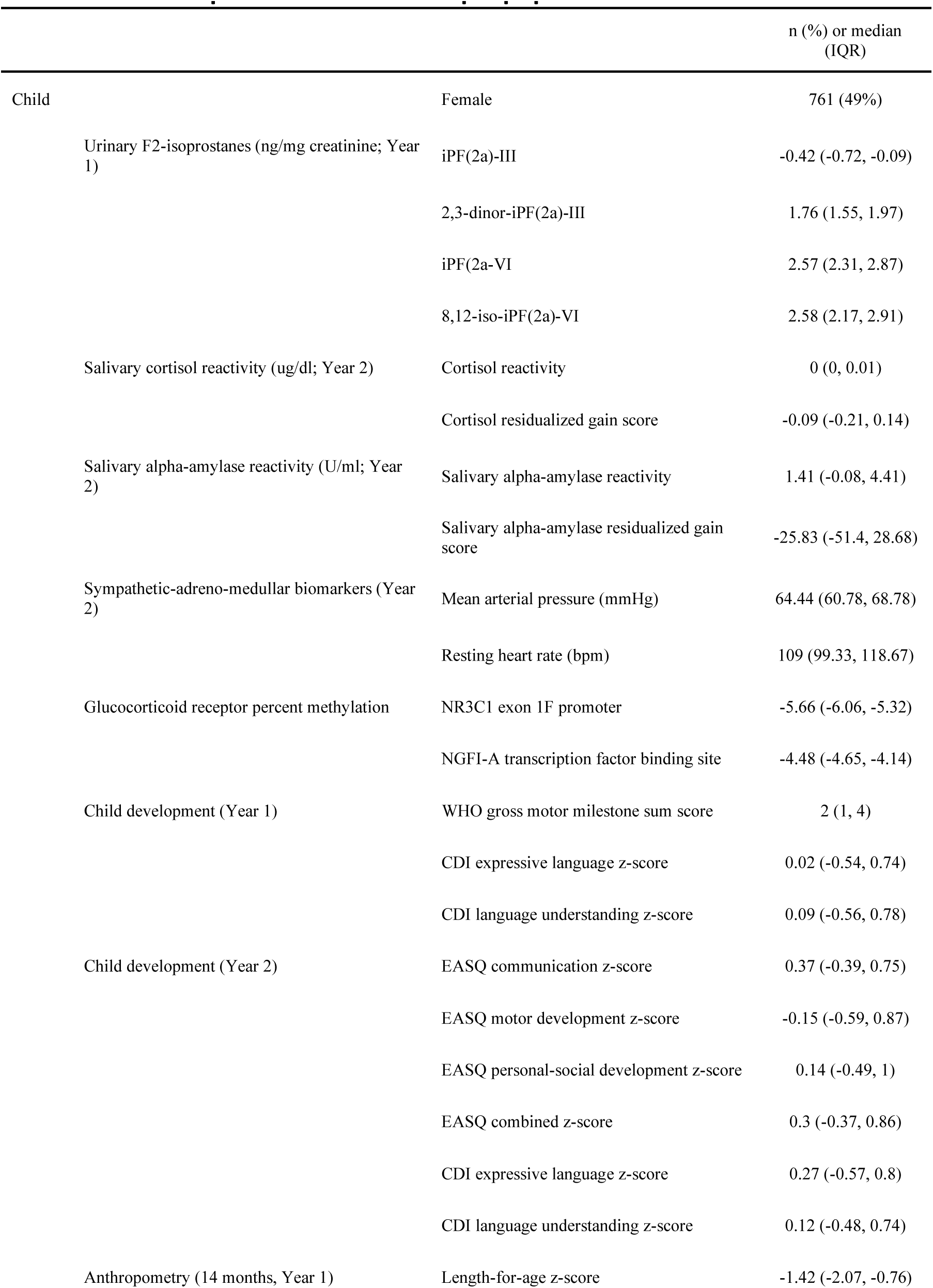
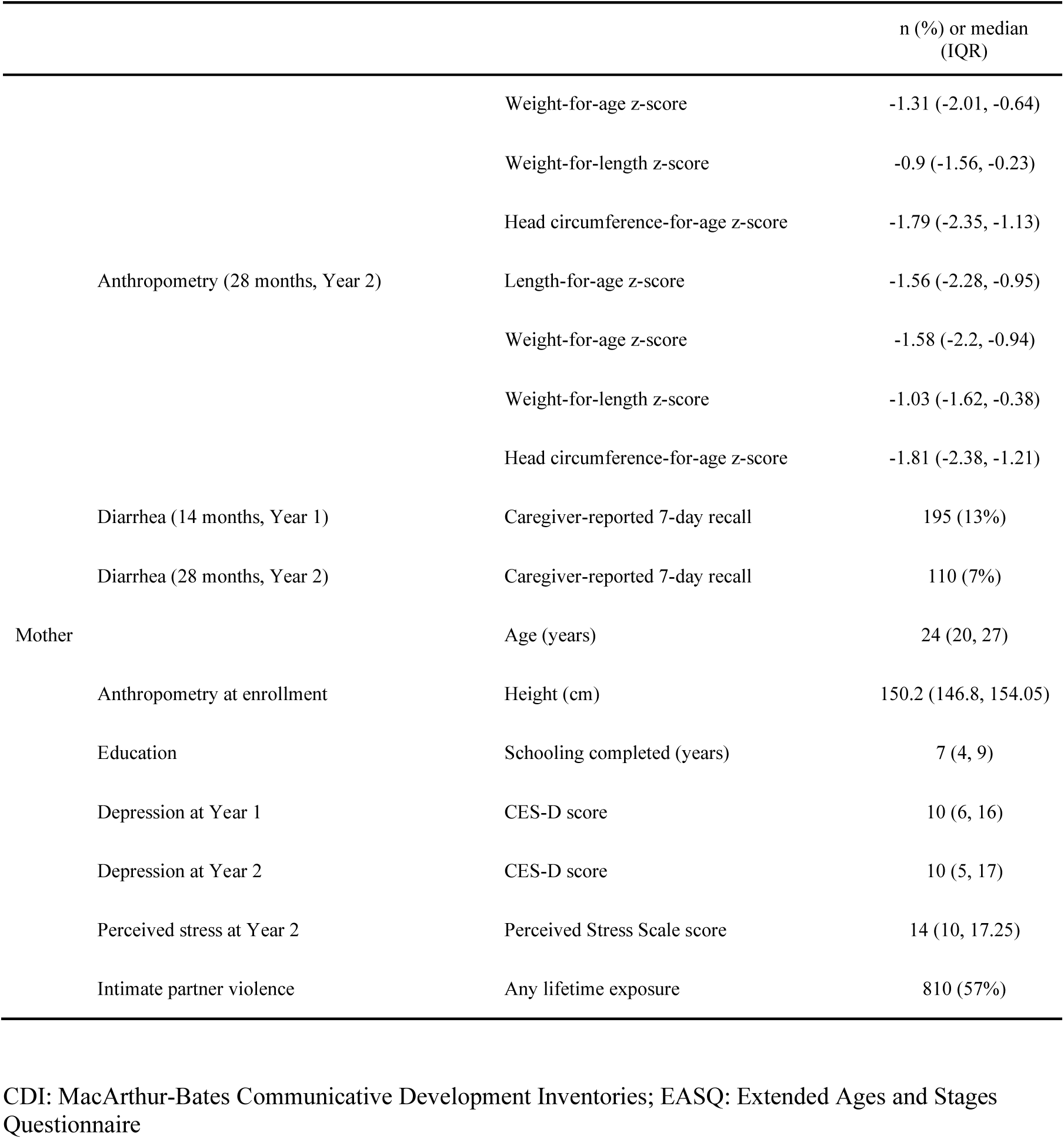
Descriptive statistics of sample population. CDI: MacArthur-Bates Communicative Development Inventories; EASQ: Extended Ages and Stages Questionnaire

We analyzed four urinary F2-isoprostane isomers separately (iPF(2a)III, 2,3-dinor-iPF(2a)III, iPF(2a)-IV, and 8,12-iso-iPF(2a)-VI), and we also used the first component of a principal components analysis of the four measures of urinary F2-isoprostanes (as these measures were correlated; P-value <0.2) to assess overall oxidative status (Jolliffe and Cadima, 2016). We assessed stress reactivity at Year 2 as the change in salivary cortisol and salivary alpha-amylase following venipuncture (see Appendix 1 for additional details). In this setting, venipuncture serves as both a physical and psychological stressor, as it involves physical discomfort as well as separation from the mother. We collected saliva pre-stressor, five minutes post-stressor, and 20 minutes post-stressor. We measured cortisol pre-and 20 minutes post-stressor, and measured alpha-amylase pre-and five minutes post-stressor (D. A. Granger et al., 2007; Douglas A. Granger et al., 2007). We calculated cortisol and alpha-amylase reactivity as the post-stressor value minus the pre-stressor value, divided by the time elapsed between samples. We recorded time of salivary biomarker assessment to account for circadian patterns of hormone production. We assessed percent methylation across the entire glucocorticoid receptor (*NR3C1*) exon 1F promoter (39 assayed CpG sites) as well as the nerve-growth factor inducing protein A (NGFI-A) transcription factor binding site, which is a specific site within the *NR3C1* exon that is associated with hippocampal glucocorticoid receptor expression (Edelman et al., 2012; Zhang et al., 2013). We measured resting heart rate and blood pressure in triplicate to ensure reliability at Year 2, where we included the median of the three measurements, and we assessed the mean arterial pressure as two times the diastolic blood pressure, plus the systolic blood pressure, divided by three (Curtis et al., 2015; Fagan et al., 1988; Rehman and Nelson, 2022). We log-transformed F2-isoprostane, cortisol, salivary alpha-amylase, and glucocorticoid receptor methylation distributions to account for skewness. We assessed child stimulation in the home through family care indicator (FCI) score as a potential effect measure modifier, which is based on Home Observations for Measurement of the Environment (Hamadani et al., 2010). The FCI includes subscales for play activities, variety of play materials, sources of play materials, books, magazines, and newspapers, and each of these subscales have demonstrated reliability for children in Bangladesh (Hamadani et al., 2010). Additional details of laboratory methods can be found in Appendix 1.

### Assessments of Child Development

Primary outcomes included child development outcomes measured via the MacArthur-Bates Communicative Development Inventories (CDI) at Years 1 and 2, the WHO gross motor milestones module at Year 1, and the Extended Ages and Stages Questionnaire (EASQ) at Year 2. Details of child development measures have previously been published and can be found in Appendix 2 (Tofail et al., 2018).

The CDI includes assessment of language expression and comprehension. WHO motor milestones include six indicators of motor development– sitting without support, hands-and-knees crawling, standing with assistance, walking with assistance, standing alone, and walking alone. Motor milestone attainment was analyzed as a sum score of the 2^nd^, 4^th^, 5^th^, and 6^th^ milestones (1^st^ and 3^rd^ milestones excluded due to a high proportion of children skipping over attainment of these indicators) to assess cumulative attainment as well as through a time-to-event analysis to assess the rate of attaining each milestone, which is consistent with previous analyses of this measure of development (Tofail et al., 2018). The EASQ has five domains, but only three were used in this study due to field-work constraints: child communication, gross motor development, and personal-social development. We also generated a combined EASQ score (Tofail et al., 2018). We age-standardized both CDI and EASQ scores using the control group as the standard population in 2-month age bins using standard techniques (Tofail et al., 2018).

### Analysis

We used R (version 4.1.1) to conduct observational analyses nested within a randomized controlled trial in accordance with a pre-registered analysis plan (https://osf.io/hzb6m/) (Lin et al., 2021a). We evaluated the association between each exposure of interest (e.g., stress reactivity at Year 2) and each outcome of interest (e.g., CDI comprehension score at Year 2) independently (Table 2), as each association potentially required its own, unique set of adjustment covariates to reduce confounding.

**Table 2.** Study hypotheses, exposures, and outcomes. CDI: MacArthur-Bates Communicative Development Inventories; EASQ: Extended Ages and Stages Questionnaire

| **Exposure Biological Domain and Biomarkers** | **Outcomes** | **Hypothesis** |
| --- | --- | --- |
| *HPA axis:* Cortisol via concentrations pre-stressor, post-stressor, and reactivity at Year 2 | CDI and EASQ at Year 2 | Cortisol concentrations and reactivity are positive correlated with child development |
| *HPA axis:* Glucocorticoid receptor methylation via percentage methylation at NGFI-A transcription factor binding site and mean overall glucocorticoid receptor methylation at Year 2 | CDI and EASQ at Year 2 | Glucocorticoid receptor methylation is negatively correlated with child development |
| *SAM axis:* Salivary alpha-amylase (concentrations pre-stressor, post-stressor, and reactivity), mean arterial pressure and resting heart rate at Year 2 | CDI and EASQ at Year 2 | SAM axis biomarkers are negatively associated with child development |
| *Oxidative status:* Individual urinary F2 isoprostane isomers and combined score at Year 1 | WHO motor milestones and CDI at Year 1; CDI and EASQ at Year 2 | Oxidative status is inversely associated with child development |

We used natural smoothing splines to accommodate potential nonlinearity and summarized mean developmental outcomes across stress biomarker distributions after controlling for potential confounders and covariates of interest in accordance with our pre-registered statistical analysis plan (Wood et al., 2016; Lin et al., 2021b; Hastie and Tibshirani, 1995). All adjusted analyses included child age and sex, and we screened the following covariates for potential inclusion: birth order, maternal age and education, food insecurity, household crowding, access to drinking water, household assets, prior growth, treatment arm, month of assessment, assessment time, and maternal depression, stress, and lifetime exposure to any type of intimate partner violence. Additional information regarding covariate screening and inclusion can be found in Appendix 3. We then plotted these general additive model curves along with simultaneous confidence intervals (Nychka, 1988). The primary contrast was the difference in the mean outcome at the 75th and 25th percentile of each exposure level after adjusting for relevant covariates, which we describe as “adjusted difference” hereafter (Lin et al., 2021b). We assessed potential modification of the association between stress biomarkers and development outcome by FCI score at Year 1 for outcomes assessed at Years 1 and 2 and Year 2 for outcomes assessed at Year 2.

As these observational analyses were exploratory in nature, interpretations included both the strength of associations between individual biomarkers as well as the consistency of the direction of these associations across related biomarker groups. While typical corrections for false discovery rate aim to determine the probability of an individual result being due to random variation by adjusting for the number of repeated tests, we aimed to evaluate whether multiple measures of a similar exposure-outcome domain (e.g., salivary cortisol and child development) indicated an underlying association. For example, if we found that a domain of exposure-outcome associations (e.g. oxidative status and subsequent development) did not indicate a consistent direction of associations (i.e. some positive correlations and some negative correlations), but individual measures indicated statistically significant associations (e.g. iPF(2a)-VI at Year 1 and EASQ personal-social score at Year 2), we concluded that these individual results may be spurious associations due to repeated testing. On the other hand, if we found a consistent direction of associations (e.g., point estimates consistently indicating positive correlations) we concluded that these observed estimates might reflect a true association between the domain of exposures and outcomes. In addition, we corrected for repeated testing to evaluate the robustness of individual associations using the Benjamini-Hochberg procedure (Benjamini et al., 2001; Ferreira, 2007).

### Ethics

Primary caregivers of all children provided written informed consent prior to enrollment. Human subjects protection committees at International Centre for Diarrhoeal Disease Research, Bangladesh (icddr,b), the University of California, Berkeley, and Stanford University approved the study protocols. Investigators registered the parent trial at ClinicalTrials.gov (NCT01590095) and a safety monitoring committee convened by icddr,b oversaw the study.

## RESULTS

We analyzed data from 684 children at Year 1 (median age 14 months) and 1,449 children at Year 2 (median age 28 months) for this study (Figure 1). The children had a median length-for-age z-score (LAZ) of -1.42 (IQR -2.07, -0.76) and a diarrhea prevalence of 13% at Year 1 and a median LAZ of -1.56 (IQR -2.28, -0.95) and a diarrhea prevalence of 7% at Year 2 (Table 1). Mothers of children in the sample had a median educational attainment of 7 years (IQR 4, 9) and a 57% prevalence of having experienced any type of intimate partner violence at some point in their lifetime. The children had a median cortisol reactivity of 0 (IQR 0.00, 0.01) ug/dl/min and a median salivary alpha-amylase reactivity of 1.41 (IQR -0.08, 4.41) U/ml/min (Table 1).

**Figure 1.**
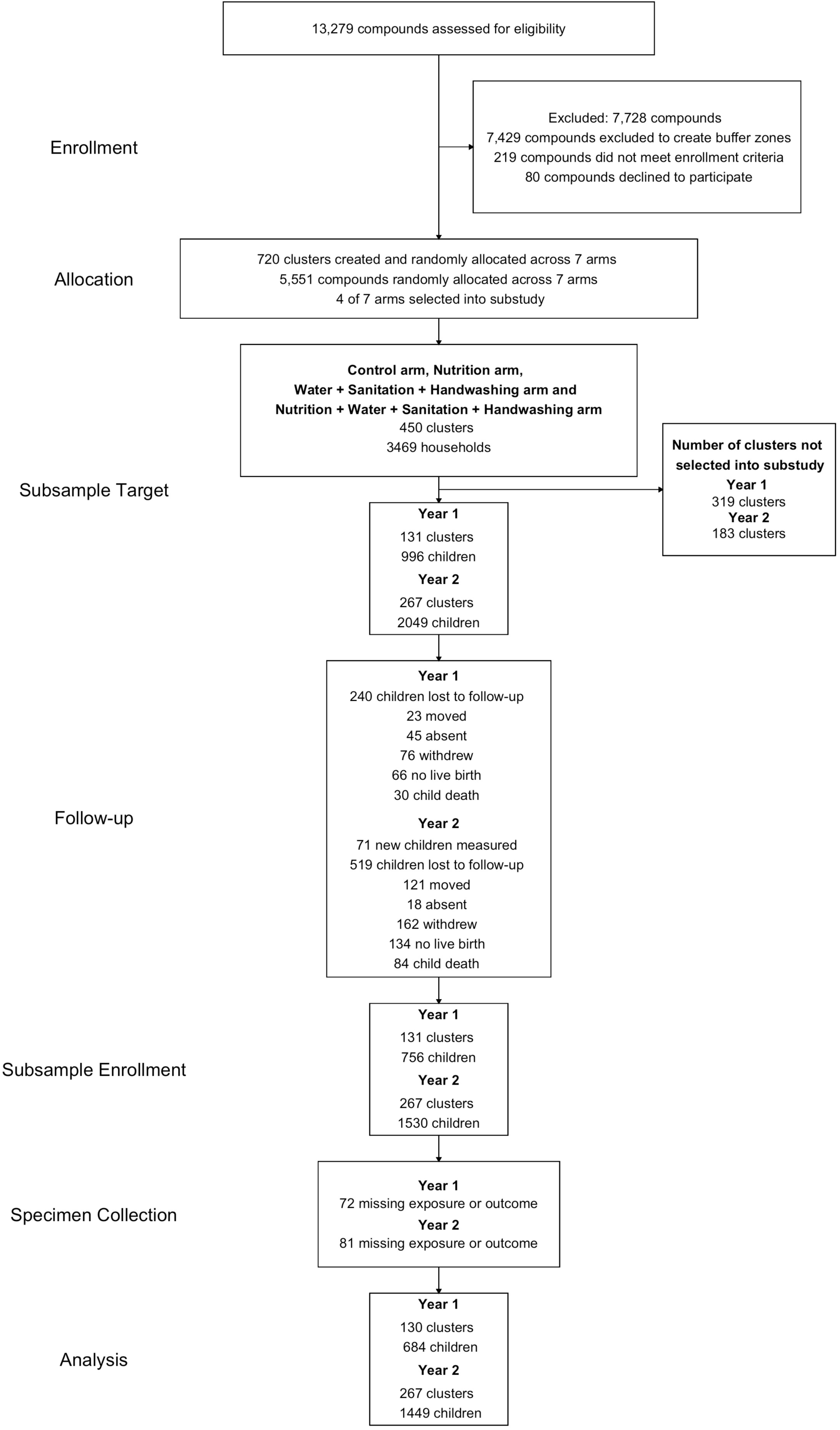
Participant enrollment, follow-up, and analysis.

We observed 135 contrasts (excluding subgroup analyses) of the differences in child development outcomes at the 75^th^ and 25^th^ percentiles of stress biomarkers across three domains of stress (HPA axis, SAM axis, and oxidative status). We found that markers of HPA axis activity (cortisol and glucocorticoid receptor methylation) were associated with child development in five out of 30 contrasts (17%; Table 3), markers of SAM activity (salivary alpha-amylase, heart rate, and blood pressure) were associated with child development in 1 out of 30 contrasts (3%; Table 4), and markers of oxidative status (F2-isoprostanes) were associated with child development in 3 out of 75 contrasts (4%, Tables 5 and 6). The proportion of significant results for HPA axis biomarkers was greater than we could expect due to random variation alone (5%; *α* = 0.05), but the proportion of significant results for SAM axis and oxidative status biomarkers were less than we would expect due to random variation, leading us to believe that significant associations with SAM axis and oxidative status biomarkers may be spurious due to repeated testing. No observed individual associations were statistically significant following false discovery rate correction for multiple testing.

**Table 3.**
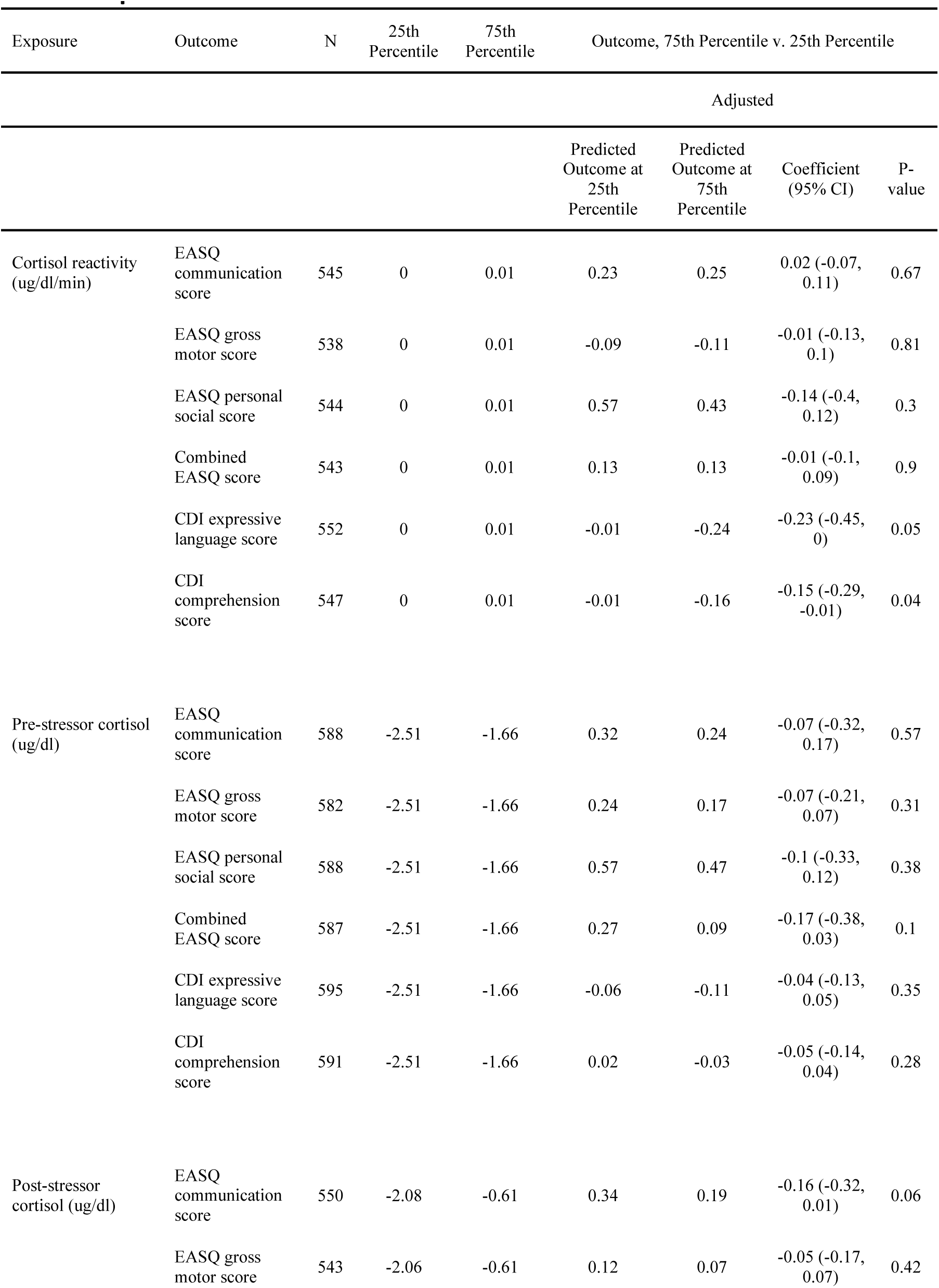
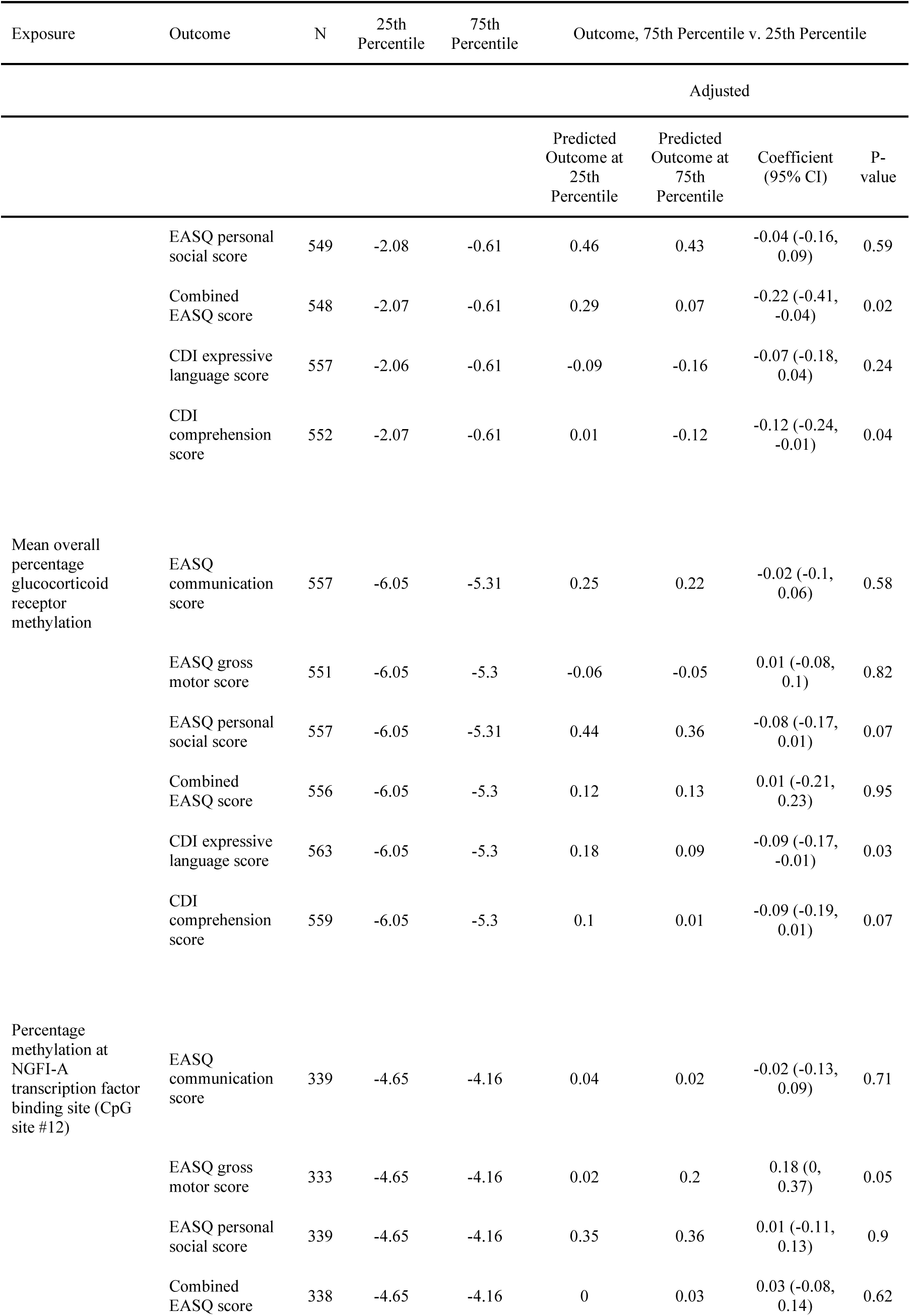
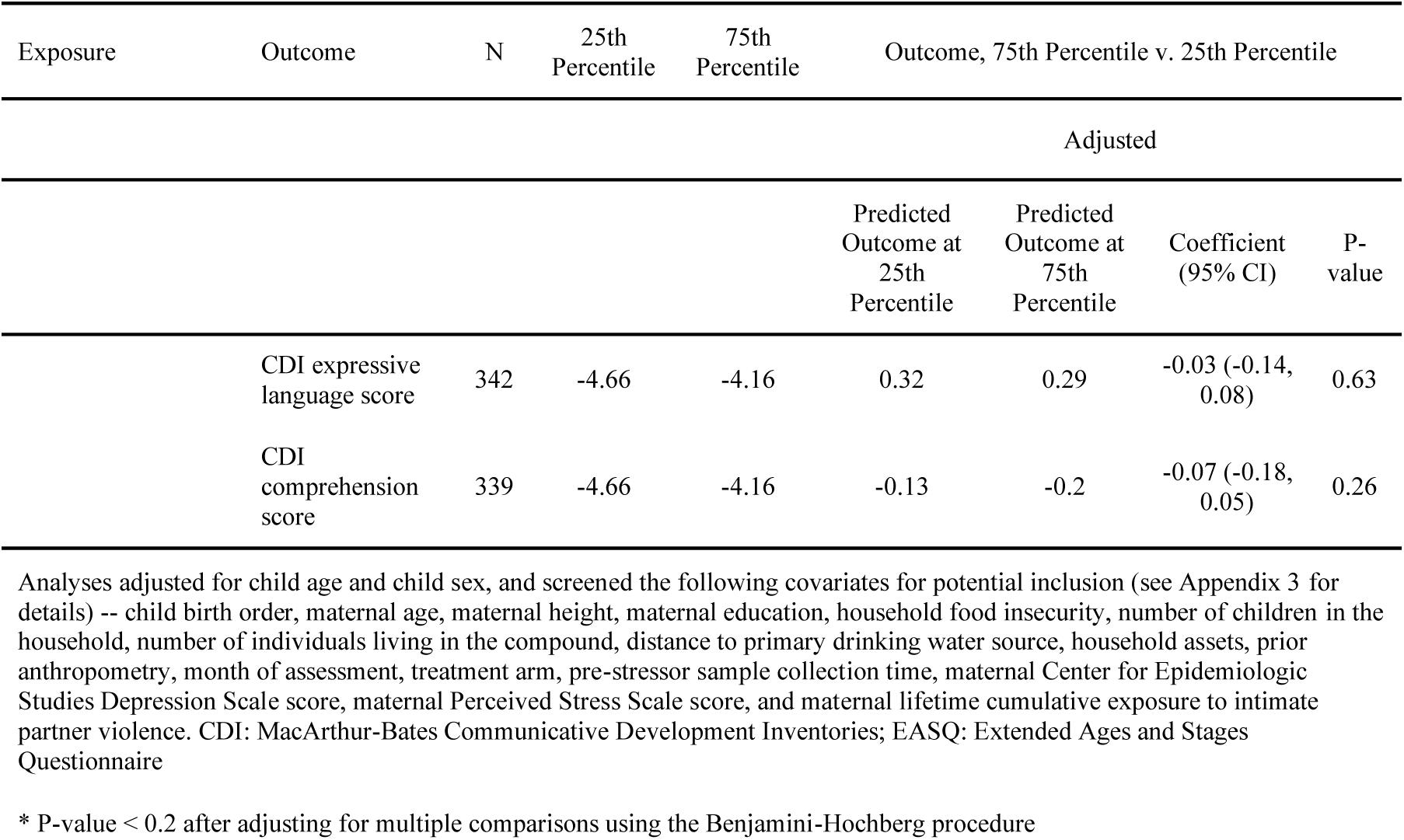
Hypothalamic-pituitary-adrenal axis biomarkers and child development at Year 2. CDI: MacArthur-Bates Communicative Development Inventories; EASQ: Extended Ages and Stages Questionnaire

**Table 4.**
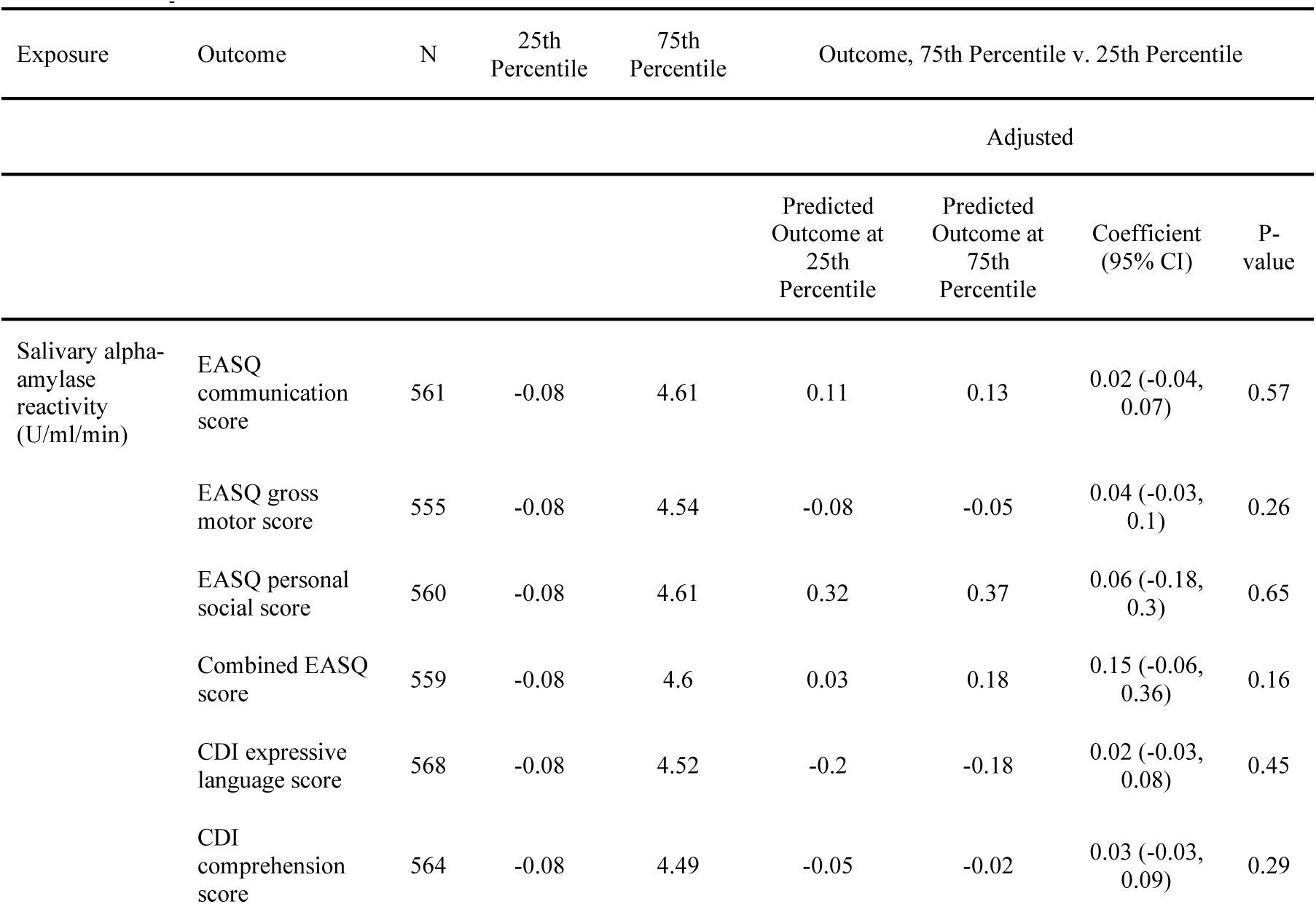
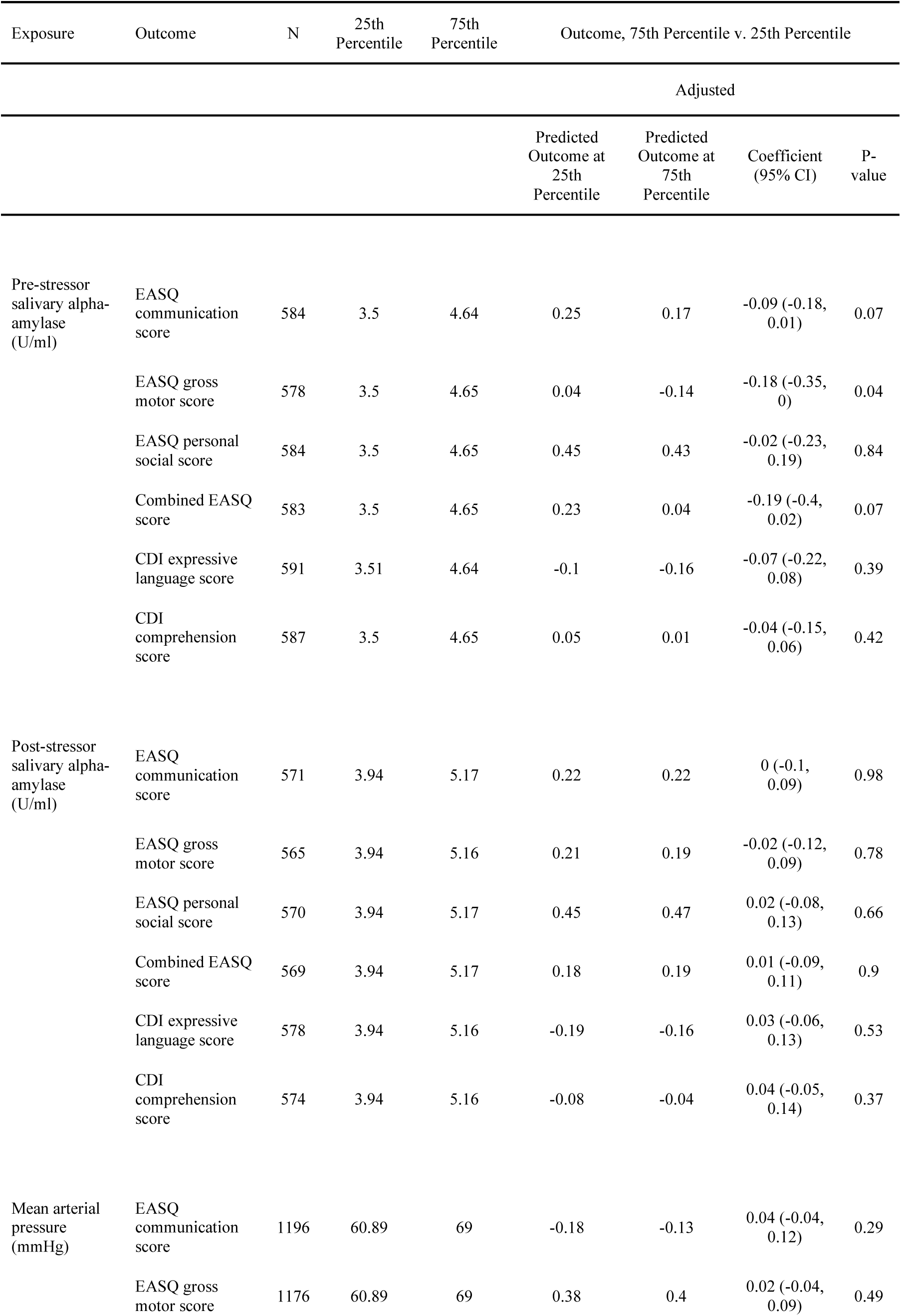
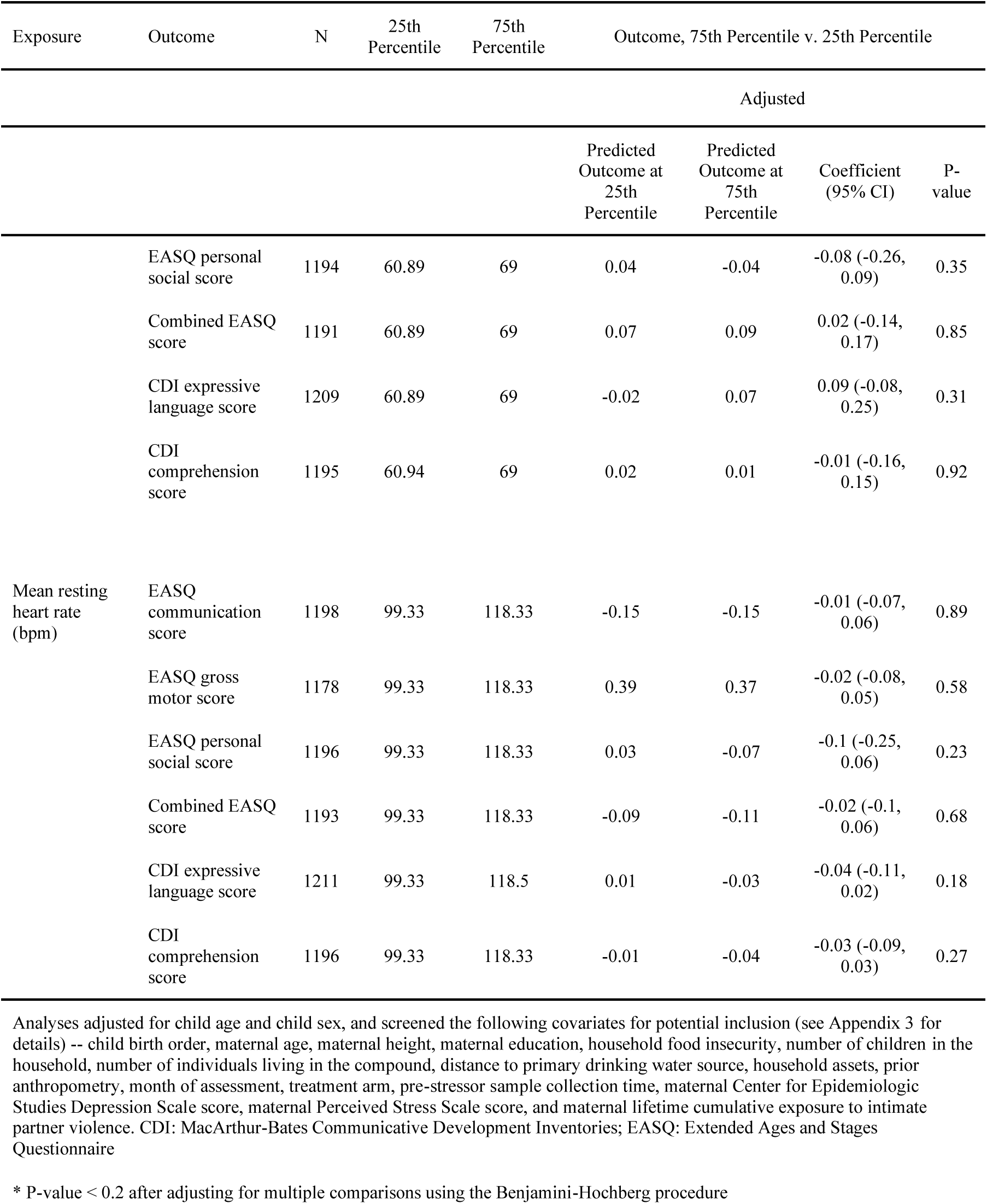
Sympathetic adrenomedullary axis biomarkers and child development at Year 2. CDI: MacArthur-Bates Communicative Development Inventories; EASQ: Extended Ages and Stages Questionnaire

**Table 5.** Urinary isoprostanes and child development at Year 1. CDI: MacArthur-Bates Communicative Development Inventories

| Exposure | Outcome | N | 25th Percentile | 75th Percentile | Outcome, 75th Percentile v. 25th Percentile |  |  |  |
| --- | --- | --- | --- | --- | --- | --- | --- | --- |
| Adjusted |  |  |  |  |  |  |  |  |
|  |  |  |  |  | Predicted Outcome at 25th Percentile | Predicted Outcome at 75th Percentile | Coefficient (95% CI) | P-value |
| IPF(2a)-III (ng/mg creatinine) | Sum of 2nd, 4th, 5th, and 6th WHO motor milestones | 571 | -0.73 | -0.09 | 1.84 | 1.79 | -0.05 (-0.37, 0.27) | 0.77 |
|  | CDI expressive language Z-score | 672 | -0.72 | -0.09 | 0.26 | 0.31 | 0.05 (-0.1, 0.2) | 0.52 |
|  | CDI comprehension Z-score | 576 | -0.73 | -0.09 | 0.37 | 0.43 | 0.06 (-0.04, 0.16) | 0.24 |
| 2,3-dinor-iPF(2a)-III (ng/mg creatinine) | Sum of 2nd, 4th, 5th, and 6th WHO motor milestones | 571 | 1.54 | 1.97 | 1.81 | 2.09 | 0.27 (0.04, 0.51) | 0.02 |
|  | CDI expressive language Z-score | 672 | 1.55 | 1.97 | 0.25 | 0.27 | 0.01 (-0.09, 0.11) | 0.8 |
|  | CDI comprehension Z-score | 576 | 1.55 | 1.97 | 0.35 | 0.36 | 0.01 (-0.11, 0.12) | 0.89 |
| iPF(2a)-VI (ng/mg creatinine) | Sum of 2nd, 4th, 5th, and 6th WHO motor milestones | 571 | 2.3 | 2.87 | 1.92 | 1.92 | 0 (-0.15, 0.15) | 0.98 |
|  | CDI expressive language Z-score | 672 | 2.31 | 2.87 | 0.2 | 0.22 | 0.02 (-0.2, 0.24) | 0.86 |
|  | CDI comprehension Z-score | 576 | 2.3 | 2.87 | 0.38 | 0.43 | 0.05 (-0.08, 0.18) | 0.43 |
| 8,12-iso-iPF(2a)-VI (ng/mg creatinine) | Sum of 2nd, 4th, 5th, and 6th WHO motor milestones | 571 | 2.17 | 2.91 | 1.9 | 1.91 | 0.01 (-0.17, 0.19) | 0.94 |
|  | CDI expressive language Z-score | 672 | 2.17 | 2.92 | 0.19 | 0.36 | 0.17 (-0.03, 0.38) | 0.09 |
|  | CDI comprehension Z-score | 576 | 2.18 | 2.91 | 0.29 | 0.44 | 0.15 (0.04, 0.27) | 0.01 |
| Adjusted |  |  |  |  |  |  |  |  |
|  |  |  |  |  | Predicted Outcome at 25th Percentile | Predicted Outcome at 75th Percentile | Coefficient (95% CI) | P-value |
| Combined urinary oxidative status score | Sum of 2nd, 4th, 5th, and 6th WHO motor milestones | 571 | 2.48 | 3.54 | 1.88 | 1.96 | 0.09 (-0.11, 0.29) | 0.41 |
|  | CDI expressive language Z-score | 672 | 2.5 | 3.55 | 0.1 | 0.31 | 0.21 (-0.03, 0.44) | 0.08 |
|  | CDI comprehension Z-score | 576 | 2.48 | 3.54 | 0.22 | 0.4 | 0.18 (-0.08, 0.44) | 0.18 |

Analyses adjusted for child age and child sex, and screened the following covariates for potential inclusion (see Appendix 3 for details) -- child birth order, maternal age, maternal height, maternal education, household food insecurity, number of children in the household, number of individuals living in the compound, distance to primary drinking water source, household assets, prior anthropometry, month of assessment, treatment arm, pre-stressor sample collection time, maternal Center for Epidemiologic Studies Depression Scale score, maternal Perceived Stress Scale score, and maternal lifetime cumulative exposure to intimate partner violence. CDI: MacArthur-Bates Communicative Development Inventories
\* P-value < 0.2 after adjusting for multiple comparisons using the Benjamini-Hochberg procedure

**Table 6.**
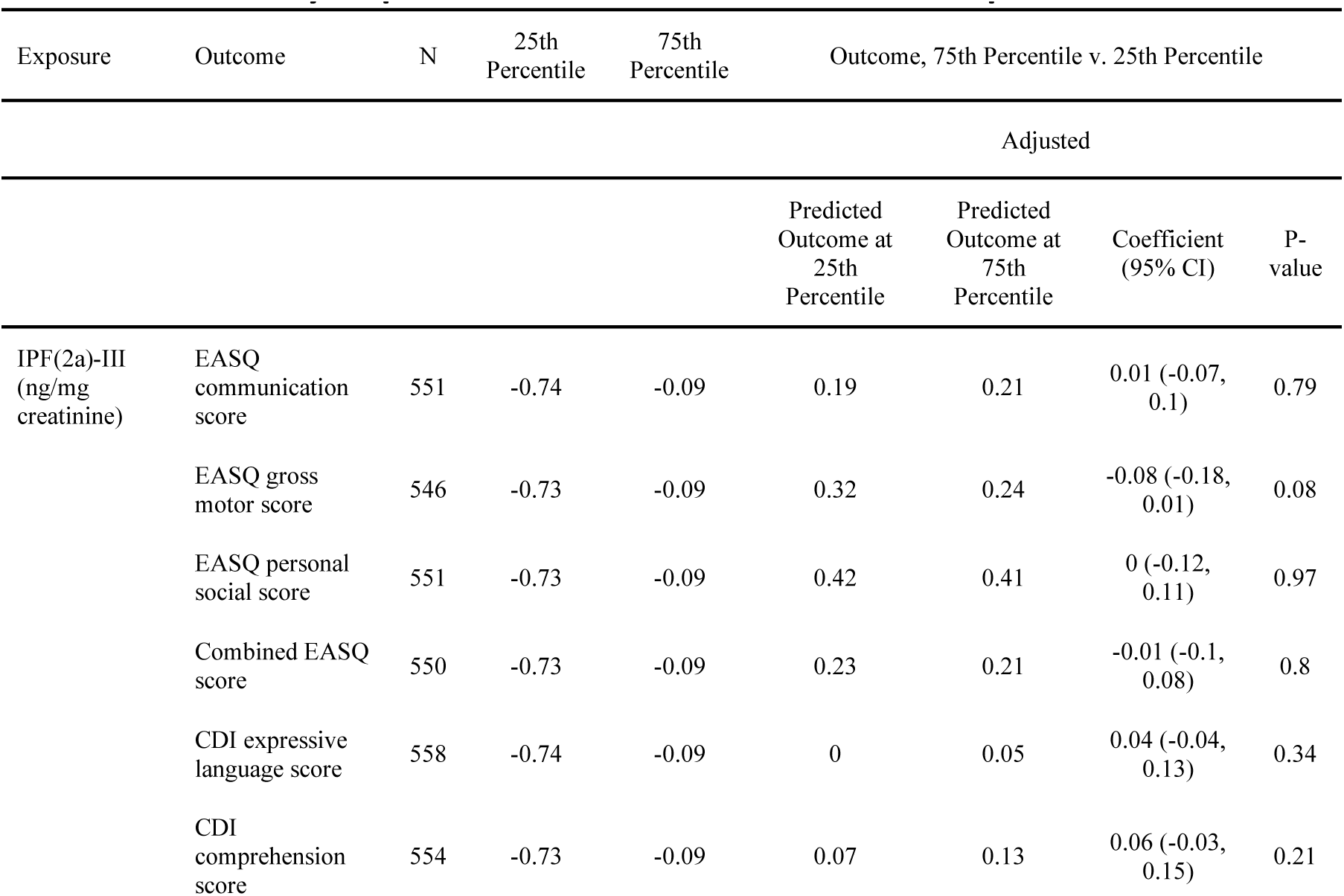
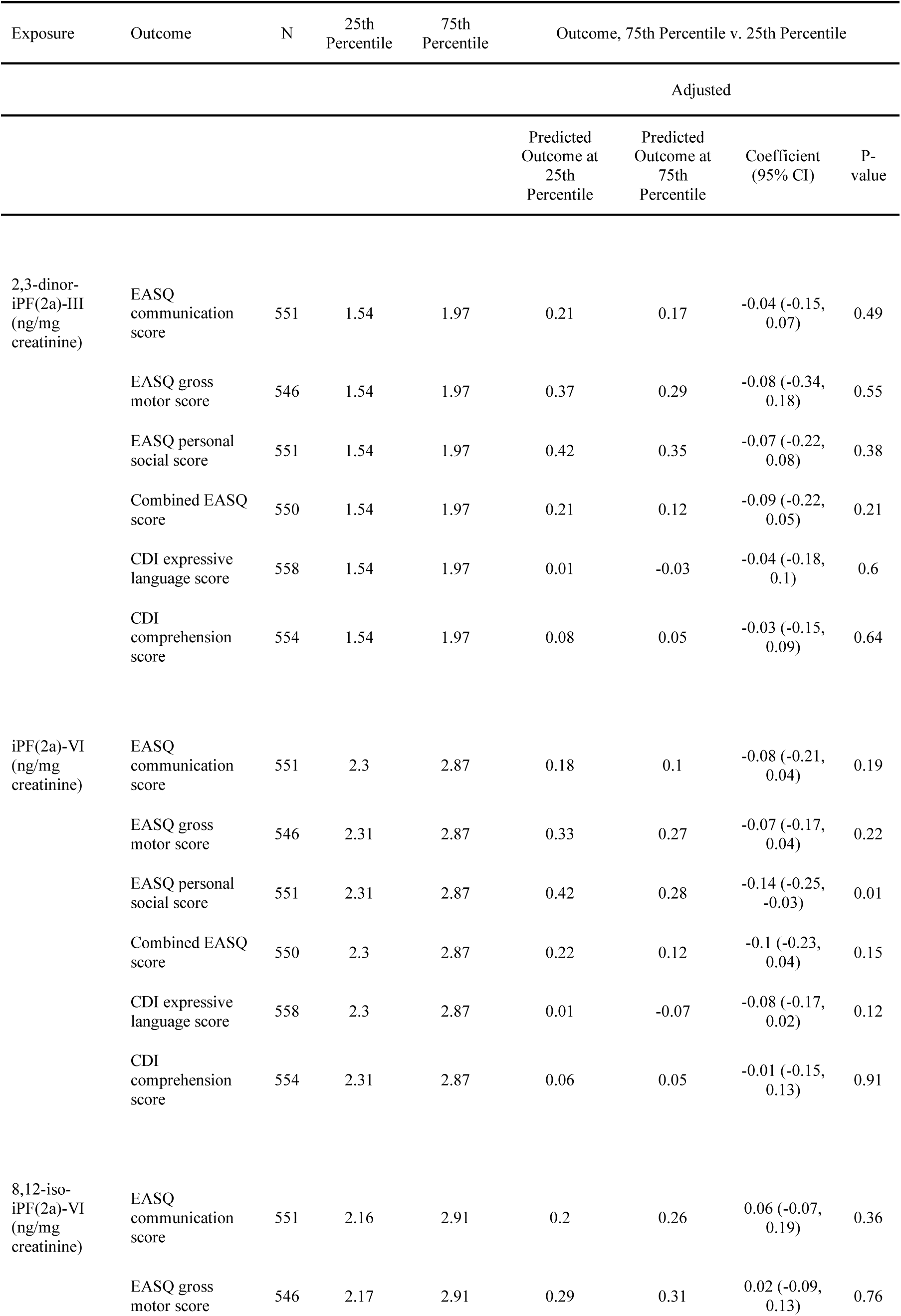
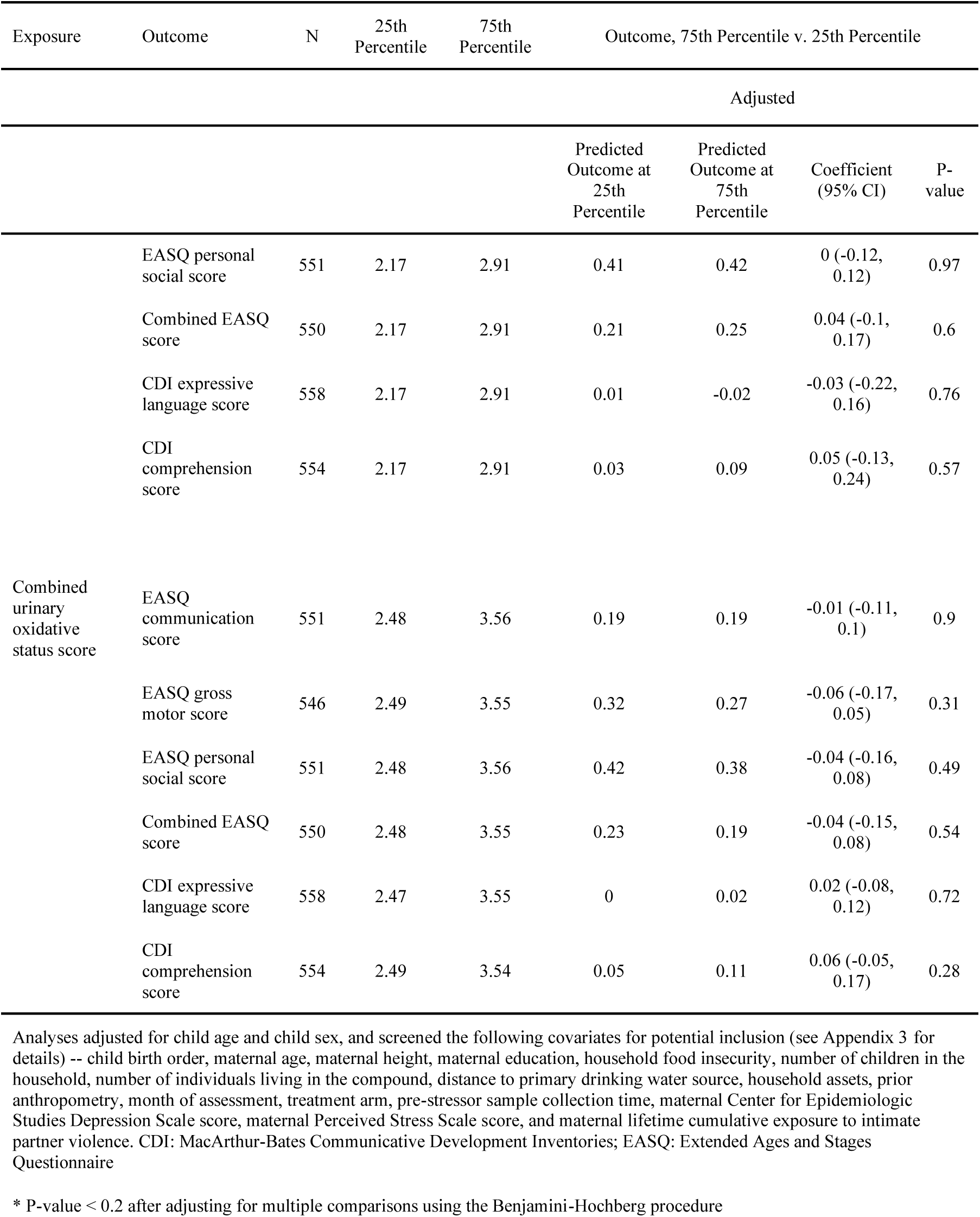
Urinary isoprostanes at Year 1 and child development at Year 2. CDI: MacArthur-Bates Communicative Development Inventories; EASQ: Extended Ages and Stages Questionnaire

### HPA axis biomarkers

We found that increased salivary cortisol production and glucocorticoid receptor methylation were associated with worse child development outcomes (Table 3, Figure 2, Figure S1). These analyses indicated that increased concurrent cortisol reactivity was associated with a lower CDI comprehension score (adjusted difference -0.15 standard deviations (SD), 95% CI (-0.29, -0.01)) and a lower CDI expression score (adjusted difference -0.23 SD, 95% CI (-0.45, 0.00)) at Year 2. In addition, we found that higher post-stressor cortisol was associated with lower combined EASQ score (adjusted difference -0.22 SD, 95% CI (-0.41, -0.04)) and lower CDI comprehension score (adjusted difference -0.12 SD, 95% CI (-0.24, -0.01)). Greater mean overall glucocorticoid receptor methylation was correlated with lower concurrent CDI expressive language score (adjusted difference -0.09 SD, 95% CI (-0.17, -0.01)), and there was a consistently negative association between overall glucocorticoid receptor methylation and concurrent measures of child development (Table 4). A greater percent methylation of transcriptor NGFI-A binding site was associated with higher EASQ gross motor score (adjusted difference 0.18 SD, 95% CI (0.00, 0.37)), but this association was not consistent across other measures of child development.

**Figure 2.**
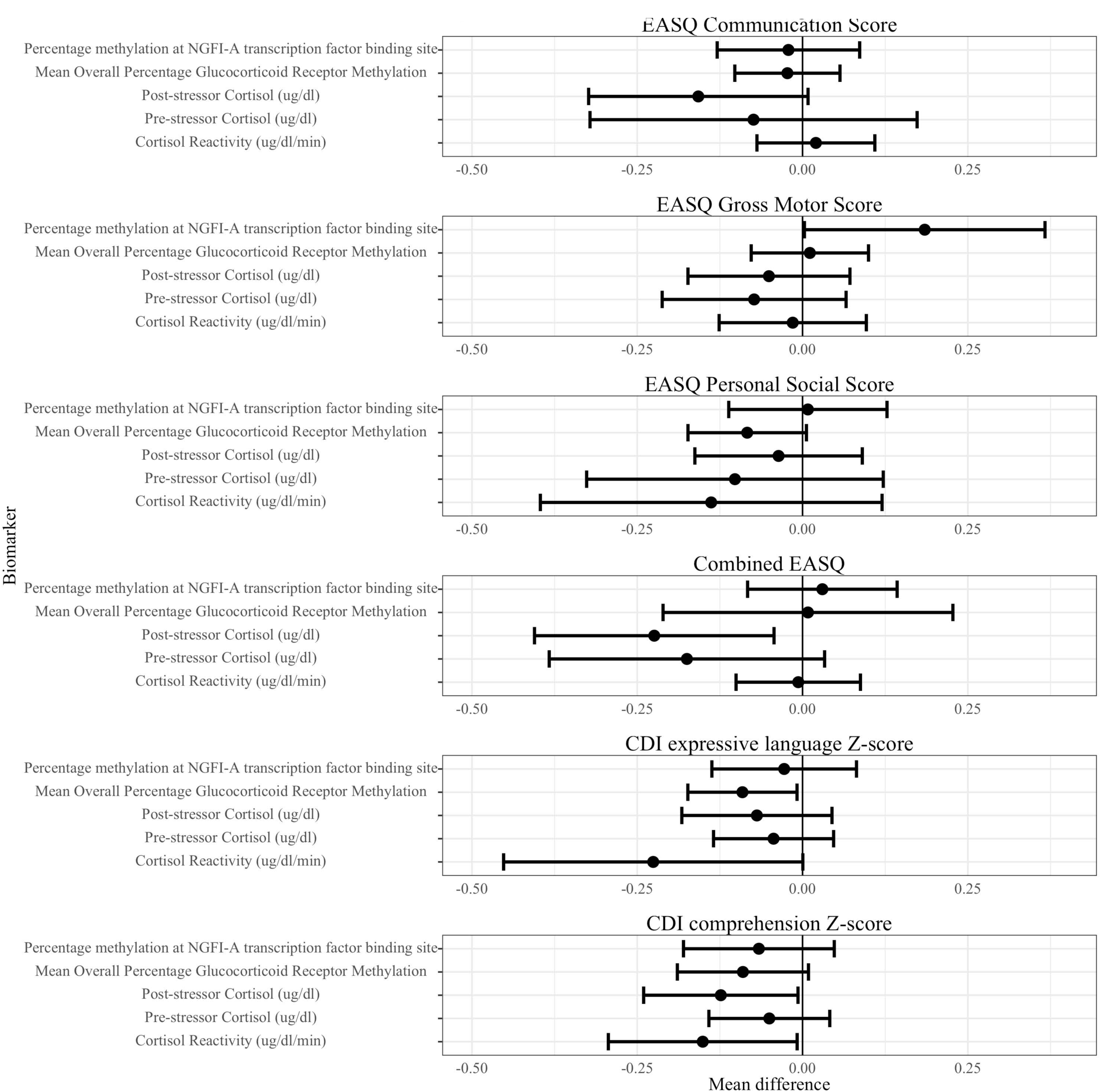
HPA axis biomarkers and child development at Year 2. HPA: Hypothalamic-pituitary-adrenal; CDI: MacArthur-Bates Communicative Development Inventories; EASQ: Extended Ages and Stages Questionnaire; sAA: salivary alpha-amylase. The difference in the mean child development outcome at the 75th and 25th percentile of each salivary stress biomarker exposure level and its 95% confidence interval after adjusting for relevant covariates.

### SAM system biomarkers

We observed that greater pre-stressor salivary alpha-amylase was associated with worse child development outcomes (Table 4, Figure 3, Figure S2). There was a significant association between pre-stressor salivary alpha-amylase and EASQ gross motor score (adjusted difference - 0.18 SD, 95% CI (-0.35, 0.00)), and the direction of this association was consistent across communication, personal social, and combined EASQ scores. We did not detect significant associations between post-stressor salivary alpha-amylase or salivary alpha-amylase reactivity and measures of development. Furthermore, we did not detect a significant association between mean arterial pressure or mean resting heart rate and any measures of concurrent development. *Oxidative status*

**Figure 3.**
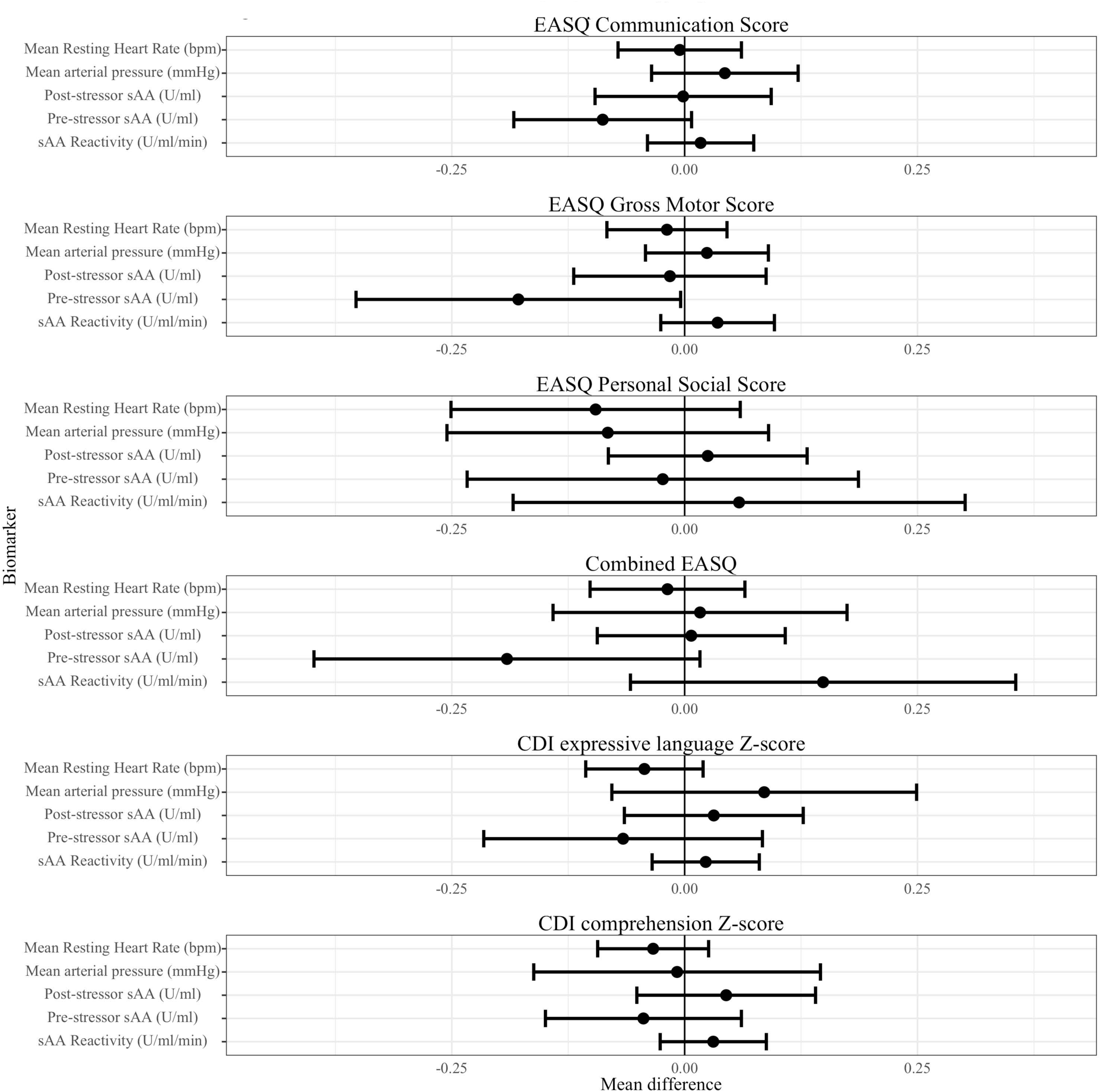
SAM axis biomarkers and child development at Year 2. SAM: Sympathetic-adrenal-medullary. CDI: MacArthur-Bates Communicative Development Inventories; EASQ: Extended Ages and Stages Questionnaire. The difference in the mean child development outcome at the 75th and 25th percentile of each measure of glucocorticoid receptor methylation and its 95% confidence interval after adjusting for relevant covariates.

We found limited evidence of an association between concurrent oxidative status and child development at Year 1 (Table 5, Figure 4, Figure S3). Increased concurrent 2,3-dinor-iPF(2a)-III (ng/mg creatinine) was associated with greater WHO motor milestones sum score (adjusted difference 0.27, 95% CI (0.04, 0.51)) as well as 8,12-iso-iPF(2a)-VI (ng/mg creatinine) and greater CDI comprehension Z-score (adjusted difference 0.15, 95% CI (0.04 SD, 0.27)). We assessed the possibility of a curvilinear association between concurrent oxidative status and child development by plotting the spline curves of these associations (Figure 5). The association between concurrent oxidative stress and CDI comprehension score largely indicated a positive correlation, while correlations of concurrent oxidative status and CDI expression and WHO sum score often depicted nonlinear associations, in which the second and third quartiles (moderate levels) of F2-isoprostanes were associated with better development outcomes relative to the first quartile.

**Figure 4.**
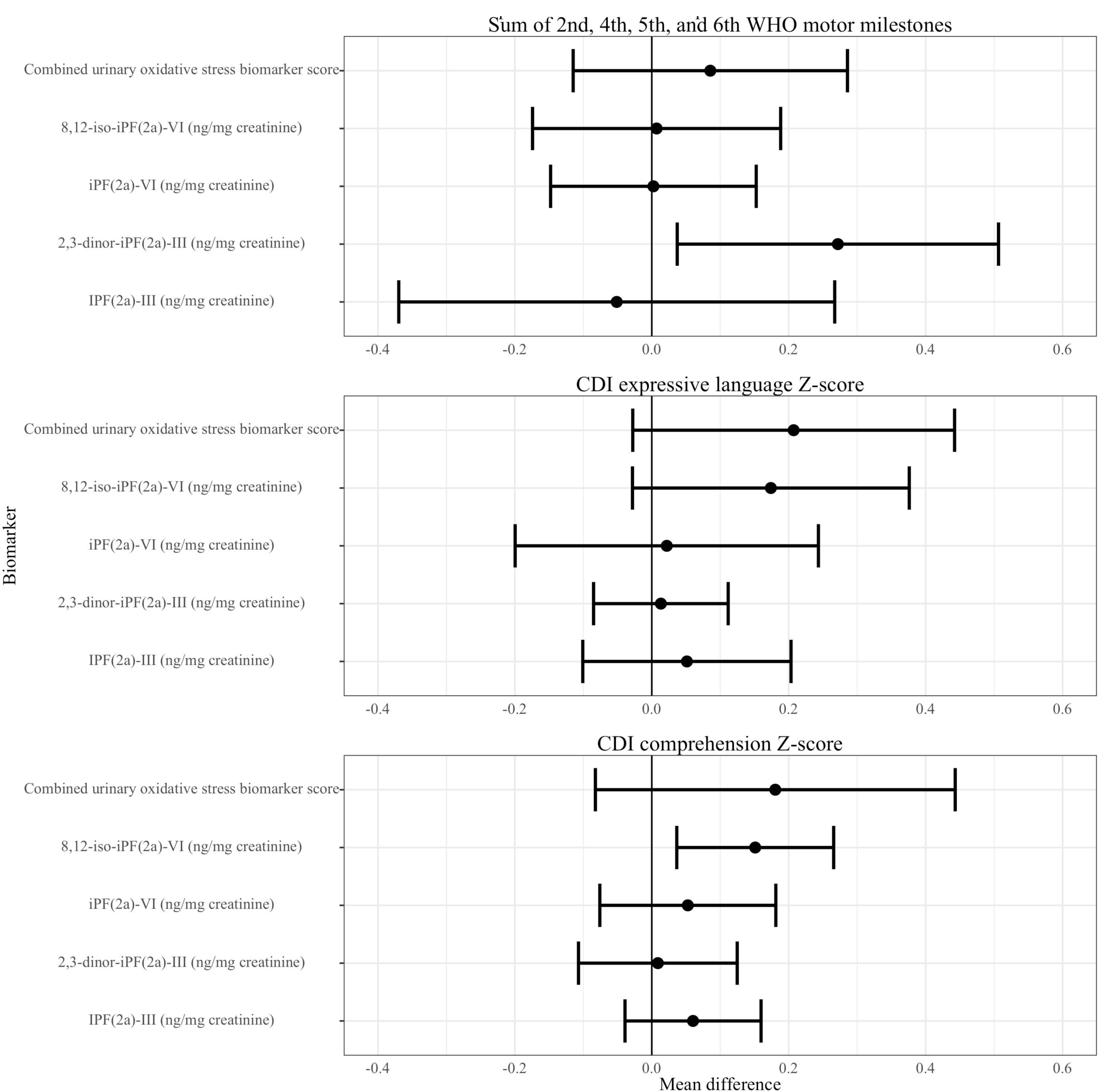
Urinary isoprostanes and child development at Year 1. CDI: MacArthur-Bates Communicative Development Inventories. The difference in the mean child development outcome at the 75th and 25th percentile of each measure of urinary isoprostanes and its 95% confidence interval after adjusting for relevant covariates.

**Figure 5.**
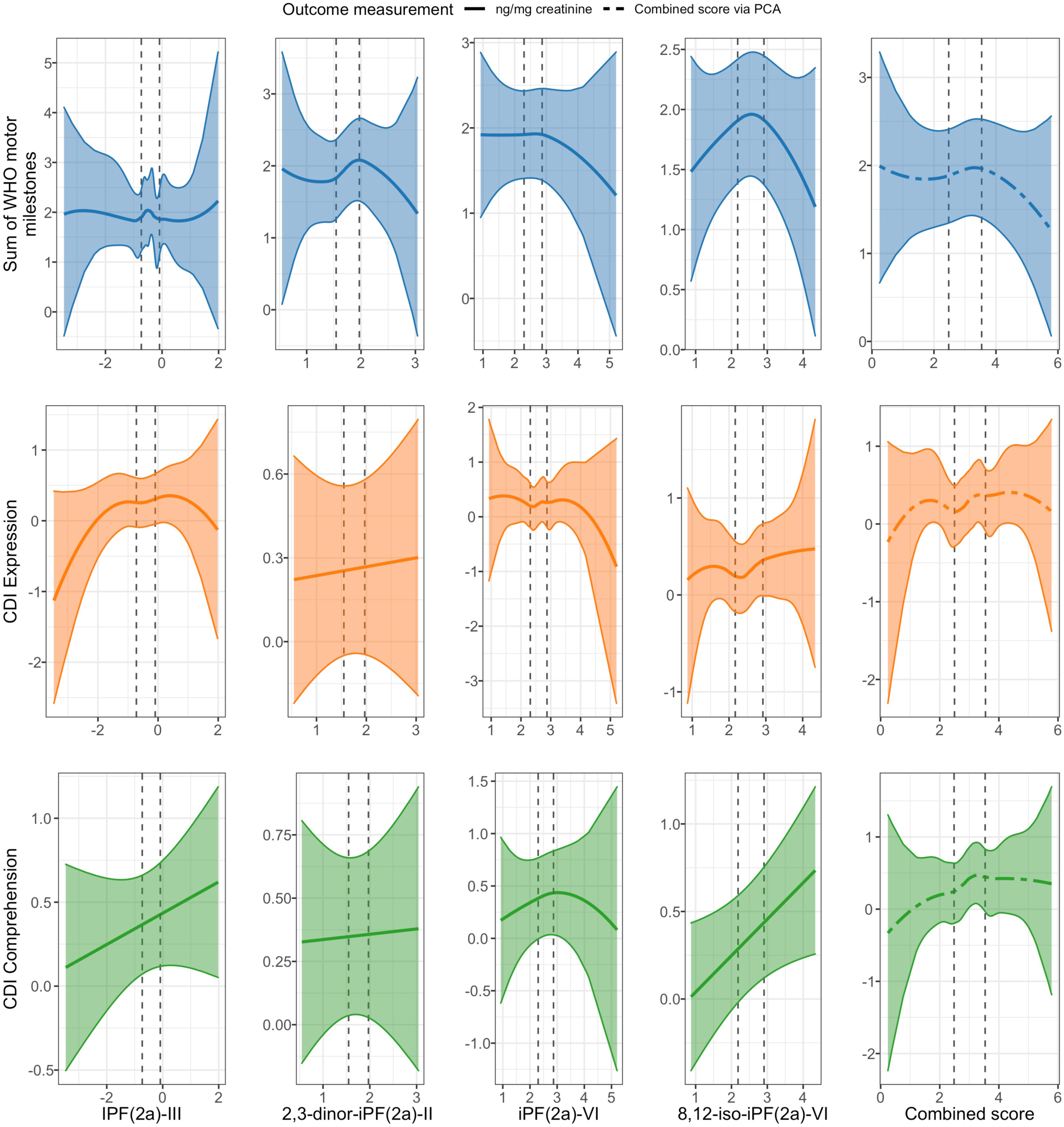
Spline curves of the associations between concurrent urinary F2 isoprostanes and child development at Year 1. CDI: MacArthur-Bates Communicative Development Inventories. Spline curves demonstrating the associations between measures of urinary isoprostanes and concurrent child development. Dotted, vertical lines represent the 25^th^ and 75^th^ percentiles of urinary isoprostane distributions.

We did not find evidence of a consistent association between measures of oxidative status at Year 1 and subsequent child development at Year 2 (Table 6, Figure 6, Figure S4). Higher levels of iPF(2a)-VI (ng/mg creatinine) were associated with lower EASQ personal-social score (adjusted difference -0.14, 95% CI (-0.25, -0.03)), but this inverse correlation was not consistent across other urinary F2-isoprostanes or measures of child development. We did not detect a significant association between any measure of oxidative status and time to attainment of any WHO motor milestone (Table 7).

**Figure 6.**
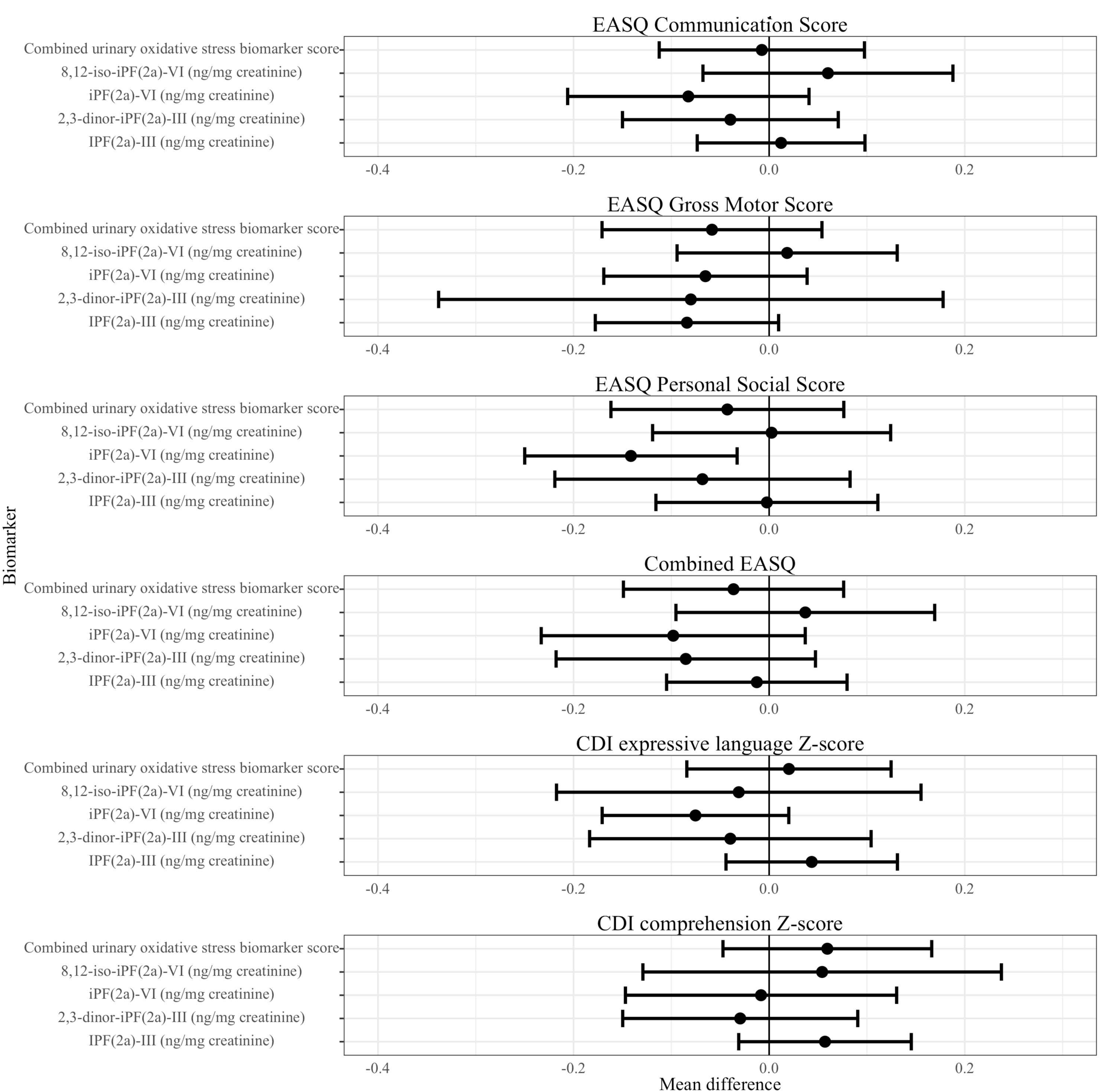
Urinary isoprostanes at Year 1 and child development at Year 2. CDI: MacArthur-Bates Communicative Development Inventories; EASQ: Extended Ages and Stages Questionnaire. The difference in the mean child development outcome at the 75th and 25th percentile of each measure of urinary isoprostanes and its 95% confidence interval after adjusting for relevant covariates.

**Table 7.**
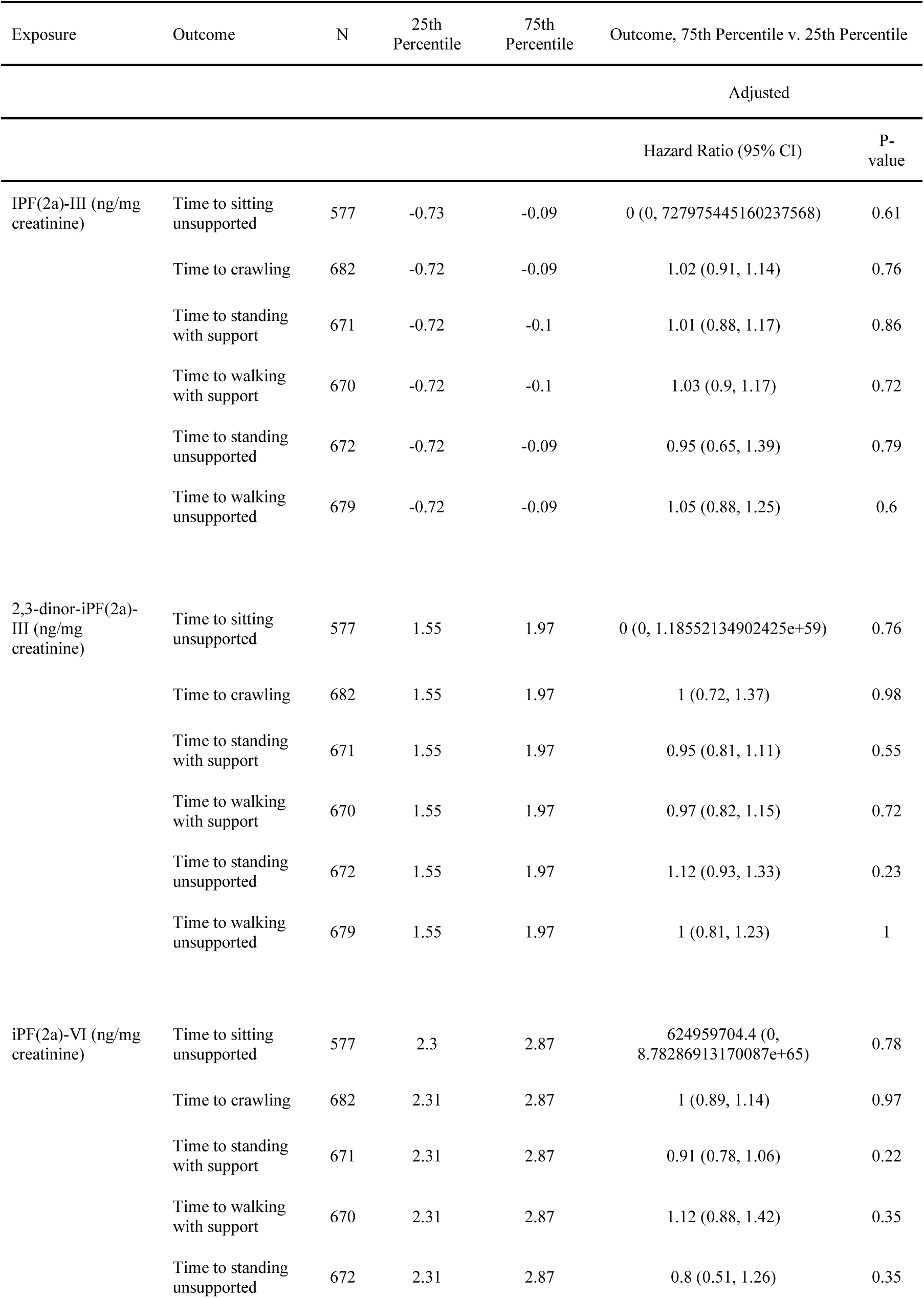
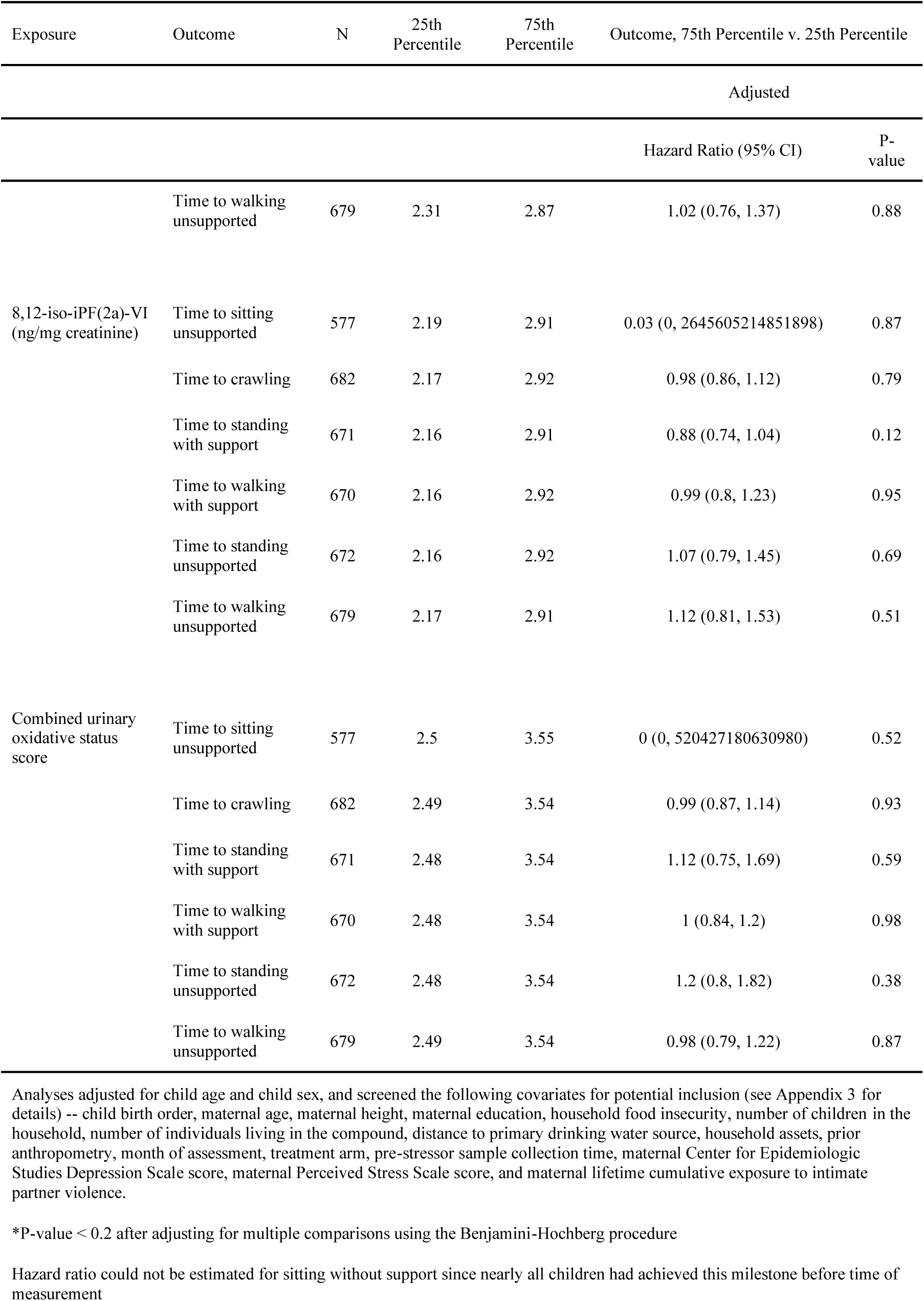
Urinary isoprostanes and time to WHO motor milestone at Year 1.

### Modification by family care indicators

We analyzed potential modification of the association between stress biomarker and development outcome by family care indicators (FCI) score at Year 1 and Year 2. The contrasted FCI scores (75th and 25th percentiles) were 9 and 5 at Year 1, and 11 and 6 at Year 2. Although we found some evidence of effect measure modification in specific exposure-outcome associations at specific timepoints, these associations were not consistent over time (e.g., FCI at Year 1 and Year 2) or across related exposure-outcome domains (Supplemental Tables 1-5).

## DISCUSSION

Our observations reveal some consistent associations between multi-level and multi-system biological signatures of exposure to chronic stress and early child development. Results suggest that individual differences in the biology of the stress response may be associated with the translation of experience and exposure into developmental consequences in early life. Yet, in this high risk (low-income, rural) developmental context in Bangladesh, using a large sample size, the magnitude of the effect was small to modest.

### HPA Axis

These findings indicate that HPA axis biomarkers, namely cortisol reactivity and overall glucocorticoid receptor methylation, are associated with developmental status, although the sensitivity of these measures is limited. This indicates that HPA axis activity may be a mechanism by which early-life adversity contributes to developmental impairment, although future studies such as mediation analyses should explore this hypothesis. While pre-stressor salivary alpha-amylase as well as moderate oxidative status showed some evidence of associations with developmental status, associations were most consistent and strongest for markers of HPA axis activity. These findings suggest that higher cortisol reactivity is associated with worse concurrent child development. This is consistent with previous studies’ findings that HPA axis hyperactivity may be related to delays in learning, memory, and neurological development (Booth et al., 2000; Franke, 2014; Grantham-McGregor et al., 2007; Lu et al., 2016; McEwen, 2011; National Scientific Council on the Developing Child, 2014). The age of the sample population has considerable implications, as early childhood is a particularly sensitive period to stress (Walker et al., 2011). In school-aged children, low or blunted (little change throughout the day) cortisol production is associated with poor developmental outcomes, and investigators have hypothesized that HPA axis hypoactivity through childhood and adulthood may be the result of early-life HPA axis hyperactivity (Gunnar and Vazquez, 2001). Follow-up evaluation of HPA axis activity in this cohort once they have reached school age may provide insights on the longer-term developmental correlates of early-life HPA axis activity.

We found that greater glucocorticoid receptor methylation was associated with worse child development outcomes. This indicates that glucocorticoid receptor hypermethylation, which is an indicator of early-life stress, may be associated with poor developmental outcomes for high-risk children in rural Bangladesh (van der Knaap et al., 2014). This is consistent with previous findings that glucocorticoid receptor methylation is positively associated with externalizing behavior and depressive symptoms in school-aged children (Cicchetti and Handley, 2017). In a previous analysis of this sample evaluating the impact of randomized assignment to interventions on child stress, we found that the control group had greater glucocorticoid receptor methylation relative to the combined nutrition, water, sanitation, and hygiene intervention group (Lin et al., 2021c). These cumulative findings indicate that glucocorticoid receptor methylation may be a pathway or marker of environmental stressors’ contribution to developmental status, but this hypothesis should be investigated further by future studies.

### SAM Axis and Oxidative Status

Although we detected a positive correlation between pre-stressor salivary alpha-amylase and child development, the lack of consistency of relationships (in terms of direction and significance) across measures of SAM axis activity and child development (3% of contrasts were significant) indicates that this association may be spurious. Similarly, although we found limited evidence that moderate oxidative status was associated with child development, the inconsistency of these correlations across measures of oxidative status and child development (4% of contrasts were significant) are quite consistent with random variation.

### Strengths and limitations

Our study evaluated the association between stress and child development using a comprehensive set of biomarkers representing the HPA axis, SAM system, and oxidative status. As each of these biomarkers reflects a unique stress response, analysis of these individual correlations between each stress biomarker and measure of child development allows for evaluation of these associations at multiple levels and multiple biological systems.

The majority of the analyses conducted were with concurrent exposure and outcome data, which does not readily enable causal inference regarding the impact of child stress on child development. We conducted observational analyses with multivariate adjustment to control for potential confounders and covariates of interest, although residual confounding may still be present. Future analyses should include a greater number of time points for observations with additional temporal separation. Our interpretations for concurrent analyses are based on the assumption that stress biomarker exposures may cause a change in child development, although it is possible that child development outcomes lead to changes in child stress neurobiology (i.e., reverse causation). In addition, assessment of child development measures for children greater than 3 years of age, such as school attendance, executive functioning, and intelligence, would enable inference of the impact of early stress on longer-term development.

The inclusion of multiple measures of both stress and development is both a strength of this study and a limitation, as multiple comparisons lead to an increased risk of Type 1 error. We aimed to account for this risk by assessing the consistency of the direction (positive vs. negative) of point estimates in each domain of exposure-outcome assessments, in addition to evaluation of each contrast’s statistical significance, although the possibility of Type 1 errors remains plausible. Furthermore, the use of corrections for false discovery, such as Benjamini-Hochberg, may be overly conservative (i.e. low power) for correlational studies of stress biomarkers and child development, where we would expect to see small to modest effect sizes. Therefore, we recommend that these inferences inform futures studies that can deliberately target and evaluate potential associations of interest.

### Conclusions

Our findings indicate a relationship between HPA axis biomarkers (cortisol and glucocorticoid receptor methylation) and developmental status of young children in Bangladesh. This indicates that HPA axis biomarkers may represent a mechanistic pathway by which early life stressors lead to subsequent development, although this hypothesis should be investigated by future studies such as mediation analyses. These associations contribute to the body of evidence that supports interventions that aim to improve child development by intervening on early-life stress, such as family-based interventions that target multiple stressors (Fisher, 2016; Pitchik et al., 2021).

## Data Availability

All data, analysis scripts, and pre-registered analysis plans are available via Open Science Framework

https://osf.io/hzb6m/

## ACKNOWLEDGMENTS

We thank the families who participated in the WASH Benefits study and the incredible icddr,b staff for their valuable contributions. This work was supported by Global Development grant [OPPGD759] from the Bill & Melinda Gates Foundation to the University of California, Berkeley and by the National Institute of Allergy and Infectious Diseases of the National Institutes of Health [grant number K01AI136885 to AL]. The content is solely the responsibility of the authors and does not necessarily represent the official views of the National Institutes of Health. icddr,b is grateful to the Governments of Bangladesh, Canada, Sweden, and the United Kingdom for providing core/unrestricted support.

