## Supplemental Materials for "Stress Biomarkers and Child Development in Young Children in Bangladesh"

*Appendix 1. Stress biomarker laboratory procedures*

F2 urinary isoprostanes: Investigators collected urine samples at Year 1 in Briggs Pediatric Sterile U-Bags and preserved samples with 0.1% thimerosal, and collected pooled aliquots over a period of five hours (protocol published elsewhere).[1] Duke University research team members evaluated four isomers of F2-isoprostanes [iPF(2α)-III; 2,3-dinor-iPF(2α)-III; iPF(2α)-VI; 8,12-iso-iPF(2α)-VI] in urine samples using liquid chromatography-tandem mass spectrometry (LC-MS/MS) and adjusted F2-isoprostanes concentrations for urine diluteness.[2]

Cortisol and salivary alpha-amylase: The caregiver was instructed not to allow their child to ingest caffeine or medicine for one hour prior to these study procedures, and we rinsed the child's mouth with drinking water 15-20 minutes prior to the stressor. Acute physical stressors (blood draw, physical exam, etc.) are often used to elicit a stress response in young children.[3,4] At Year 2, we collected one saliva sample five to eight minutes before the stressor onset, a second sample five minutes following stressor onset, and a third sample 20 minutes after the stressor onset using SalivaBio Children's Swabs (Salimetrics). The International Centre for Diarrhoeal Disease Research, Bangladesh (icddr,b) Mymensingh satellite laboratory evaluated sAA and cortisol (Salimetrics) with commercial ELISA kits. We measured both sAA and cortisol at two time points.

Glucocorticoid receptor methylation: We collected saliva samples for DNA methylation analysis in Oragene kits (OGR-575). EpigenDx (Hopkinton, MA) extracted DNA from these samples, conducted bisulfite treatment and pyrosequencing, performed PCR, and determined percent methylation.[5]

Mean arterial pressure and resting heart rate: We measured resting heart rate using a finger pulse oximeter (Nonin 9590 Onyx Vantage) and blood pressure (systolic and diastolic) using a blood pressure monitor (Omron HBP-1300) in triplicate at Year 2.

*Appendix 2. Child development outcomes*

MacArthur-Bates Communicative Development Inventories (Year 1 and 2): We administered a culturally adapted assessment of language development based on the CDI, which is parentally reported and has been validated for use in Bangladesh.[6,7]

WHO motor milestones: We assessed gross motor development using the WHO motor milestones module which uses both parental report and direct assessment.[8]

Extended Ages and Stages Questionnaire: The EASQ primarily involves parental report of child development, although it also includes child demonstration of specific behaviors.[6] We adapted this tool for use in Bangladesh by adding direct assessment to 25% of the items with the support of International Centre for Diarrhoeal Disease Research, Bangladesh (icddr,b) psychologists.[39] The EASQ has been validated to accurately capture variation in child development across different levels of socioeconomic status and at-home child stimulation in low-income countries.[9,10] We determined child age from child birth dates and verified this age, when possible, using vaccination cards.

*Appendix 3. Analysis methods and covariates*

This analysis controlled for child age, child sex, and covariates that were significantly associated with the outcome of interest in each unique analysis. We assessed the following baseline enrollment covariates – child birth order, maternal age, maternal height, maternal education, household food insecurity, number of children in the household, number of individuals living in the compound, distance to primary drinking water source, and a household asset score calculated via the first principal component of a principal components analysis. In addition, based on literature review and subject area knowledge, we identified time-varying covariates that were possibly associated with the outcomes of interest but were not likely to be caused by either the exposure or outcome of interest. These time-varying covariates included – prior length-for-age Z-score (LAZ, categorized), prior weight-for-age Z-score (WAZ, categorized), month of measurement, treatment arm (control, N+WSH), pre-stressor sample collection time, maternal Center for Epidemiologic Studies Depression (CES-D) Scale score,[11] maternal Perceived Stress Scale (PSS) score,[12] and maternal lifetime cumulative exposure to intimate partner violence as measured by the WHO Women's Health and Life Experiences Survey.[13] Additional information regarding the rationale for covariate inclusion and temporality of assessment can be found in this study's pre-registered analysis plan.[14] We prescreened each covariate meeting this criteria using a likelihood ratio test to assess potential relationships between each covariate and each outcome, where we included each covariate that yielded a p-value less than 0.2 and excluded covariates with little (<5%) variation in the study sample.
